# Cost-effectiveness of maternal vaccine and/or monoclonal antibody strategies against respiratory syncytial virus in Belgian infants

**DOI:** 10.1101/2025.10.31.25339272

**Authors:** Xiao Li, Lander Willem, Dominique Roberfroid, Joke Bilcke, Diego Castanares Zapatero, Christophe De Meester, Zhuxin Mao, Nancy Thiry, Philippe Beutels

**Affiliations:** Centre for Health Economics Research and Modelling Infectious Diseases (CHERMID), Vaccine & Infectious Disease Institute, University of Antwerp, Antwerp, Belgium; Primary and Interdisciplinary Care Antwerp (ELIZA), University of Antwerp, Antwerp, Belgium; Belgian Health Care Knowledge Centre (KCE), Brussels, Belgium

## Abstract

The respiratory syncytial virus (RSV) burden and cost-effectiveness of infant RSV immunisation was evaluated by comparing seven strategies in terms of costs and Quality-Adjusted Life Years (QALYs): no universal immunisation, year-round or seasonal maternal vaccination (MV), year-round or seasonal nirsevimab (NmAb) at birth, seasonal NmAb plus a catch-up for infants≤6-month and a combined MV+NmAb strategy. Seasonal NmAb with catch-up was most effective, while seasonal MV was least effective, but most cost-effective for healthcare at current list prices (MV €186, NmAb €778). Extensive trade-offs between NmAb and MV show at which cost per dose which strategy would be deemed cost-effective. At willingness-to-pay €35,000/QALY gained, seasonal NmAb+catch-up was preferred if NmAb<€210; otherwise, seasonal or year-round MV was preferred when MV < €220 or < €75, respectively. The combined strategy became optimal at low MV and NmAb cost levels. Besides price level, cost-effectiveness was most sensitive to RSV hospital burden.

## Introduction

Respiratory syncytial virus (RSV) is a major public health concern in young children, especially infants. It often causes serious illnesses like bronchiolitis and pneumonia, leading to hospitalisation and possibly paediatric intensive care unit (ICU) admissions. In high-income countries, RSV-related lower respiratory tract infections (LRTIs) affect about 38.5 per 1000 infants annually, with the highest hospitalisation rates seen in those aged 0–3 months ^1,2^.

A retrospective analysis of the Belgian national hospital discharges reported 8,046 RSV-related hospitalisations and 4 in-hospital deaths among infants in the calendar year 2018. Among hospitalised neonates (0–28 days old), 15.9% were referred to PICU ^3^. The associated hospitalisation incidence was 68.3 per 1,000 infants <1 year and 5 per 1,000 in children aged 1–4 years ^3^. The hospitalisation incidence in infants stands well above typical European estimates of 20–40 per 1,000 infants ^4^.

RSV is highly seasonal, with peak activity concentrated typically in the winter months, alongside other respiratory pathogens. This also leads to significant organisational and economic burdens on primary, secondary, and tertiary health care services.

RSV-related admissions occupied an estimated 20% to 40% of paediatric hospital beds during the seasonal peak in 2018 ^3^. Another Belgian regional study estimated that RSV-related hospitalisations in children under 3 years of age cost >€26 million from the perspective of the national health insurance system ^5^. In addition, RSV causes important health-related quality of life (HRQoL) losses to both infants and their parents/caregivers ^6^.

Monoclonal antibody palivizumab (Synagis®) has been available since the late 1990s to protect high-risk infants against RSV ^7^, but its use has remained limited due to high cost, monthly dosing, and narrow eligibility, prompting the development of broader, more practical immunisation alternatives. In 2022-2023, a single-dose long-acting monoclonal antibody nirsevimab (NmAb, Beyfortus®) and a maternal vaccine (MV, Abrysvo®) were licensed in several high income countries ^8,9^. Another single-dose monoclonal antibody, clesrovimab (Enflonsia®), was also approved in 2025 in the United Sates. In December 2023, Belgian’s National Immunization Technical Advisory Group (NITAG) of the Superior Health Council (SHC), recommended using either MV or NmAb for RSV prevention in infants ^10^. MV is recommended for pregnant women between 28–36 gestation weeks expecting to deliver from September to March, unless they are at risk of reduced vaccine effectiveness (e.g. immunocompromised or premature delivery), in which case NmAb is preferred.

NmAb is recommended for all infants born to unvaccinated mothers, born prematurely (<30 weeks), or born within 2 weeks of maternal vaccination. It is to be given once at birth or during a routine immunisation visit for healthy infants ≤6 months old, and in high-risk children during their first and second RSV seasons.

Both MV and NmAb were conditionally reimbursed in Belgium, subject to prior authorisation. As of May 2024, NmAb is reimbursed under specific criteria, with reimbursement for full-term infants (≥36 weeks gestation) granted temporarily until May 31, 2026 ^11^. While the list price is €777.58 per dose (50 mg or 100 mg), the actual reimbursed price remains confidential ^12^. The MV has been reimbursed since January 2025 for pregnancies with an expected delivery date during the RSV season (September–March), at a list price of €186.01 per dose ^13^.

In order to independently inform decision making in Belgium, the current study was requested by regional authorities and coordinated through the Belgian Health Care Knowledge Centre (KCE)^14^. As such, the study aimed to estimate the disease burden in children under 5 years and to make a full incremental cost-effectiveness and a budget impact analysis of RSV prevention strategies to inform reimbursement, immunisation program funding and implementation decisions.

## Results

### RSV-related disease and economic burden

By following the Belgian birth cohort over a 5-year time horizon, we estimated in the absence of universal RSV immunisation approximately 116 thousand RSV cases, including 40 thousand non-medically attended cases, 66 thousand outpatient cases, 8,638 non-ICU hospitalisations, 428 ICU admissions, and 5 deaths. The highest rates of deaths and hospital admissions occurred in infants aged 0–2 months, while this group had the fewest outpatient and non-medically attended episodes. Overall, RSV was associated with an estimated 968 undiscounted Quality-Adjusted Life Years (QALYs) lost, with over 36% due to mortality in children under 5. From the health care payer perspective, RSV incurred €43 million undiscounted medical costs, over two-thirds of which were attributable to children under 1 year (Table S.11).

### Impact of RSV interventions on burden

In our base case, the following 5 immunisation programmes were compared to no intervention and to each other through a full incremental cost-effectiveness analysis:

- **“MV”**: Year-round single-dose maternal vaccine during pregnancy
- **“NmAb”**: Year-round single-dose nirsevimab at birth
- **“MV: Sep-Mar”**: Seasonal maternal vaccine during pregnancy for infants with a due delivery date from September to March
- **“NmAb: Oct-Mar”**: Seasonal nirsevimab given at birth for infants born during the RSV season from October to March
- **“NmAb: Oct-Mar + catch-up”**: Seasonal nirsevimab (as described above) plus a catch-up programme in September for infants (≤6 months) born outside of the RSV season from April to September

In the scenario analysis, we evaluated an additional “combined” strategy defined as

- **“MV+NmAb: Oct-Mar + catch-up”**: Seasonal maternal vaccine (as described above) at 40% coverage, supplemented with NmAb for 90% of infants born during the RSV season and of which the mother was not vaccinated, and an out of season NmAb catch-up programme (as described above).

Compared to no intervention, seasonal MV and NmAb strategies prevented 18% and 45% fewer RSV cases respectively, than the equivalent year-round strategies, but they incurred substantially lower costs (Table S. 12). The seasonal plus catch-up NmAb strategy averted the largest RSV burden, preventing nearly 20 thousand cases and gaining 216 discounted QALYs, with €19 million in medical costs averted. However, when assuming list prices, this strategy incurred the highest intervention cost (€76 million at 90% coverage) due to the large target group and high price (€777.58 per dose) ^12^, whereas the seasonal MV strategy, with 40% coverage and a price of €186.01 per dose ^13^, resulted in a lower intervention cost of €5 million. As protection lasts up to 6 months and no infants over 6 months were targeted, the averted disease burden was limited to infants (<1 year).

### Baseline cost-effectiveness

As illustrated in the cost-effectiveness plane (Figure 2) from health care payers (HCP) prespective, the seasonal MV, year-round MV, and seasonal plus catch-up NmAb strategies lay on the frontier at list prices. Under the cost parity assumption (€200 per dose), the NmAb seasonal and NmAb seasonal plus catch-up strategies formed the frontier, reflecting a shift in efficiency at equal costs.

**Figure 1.**
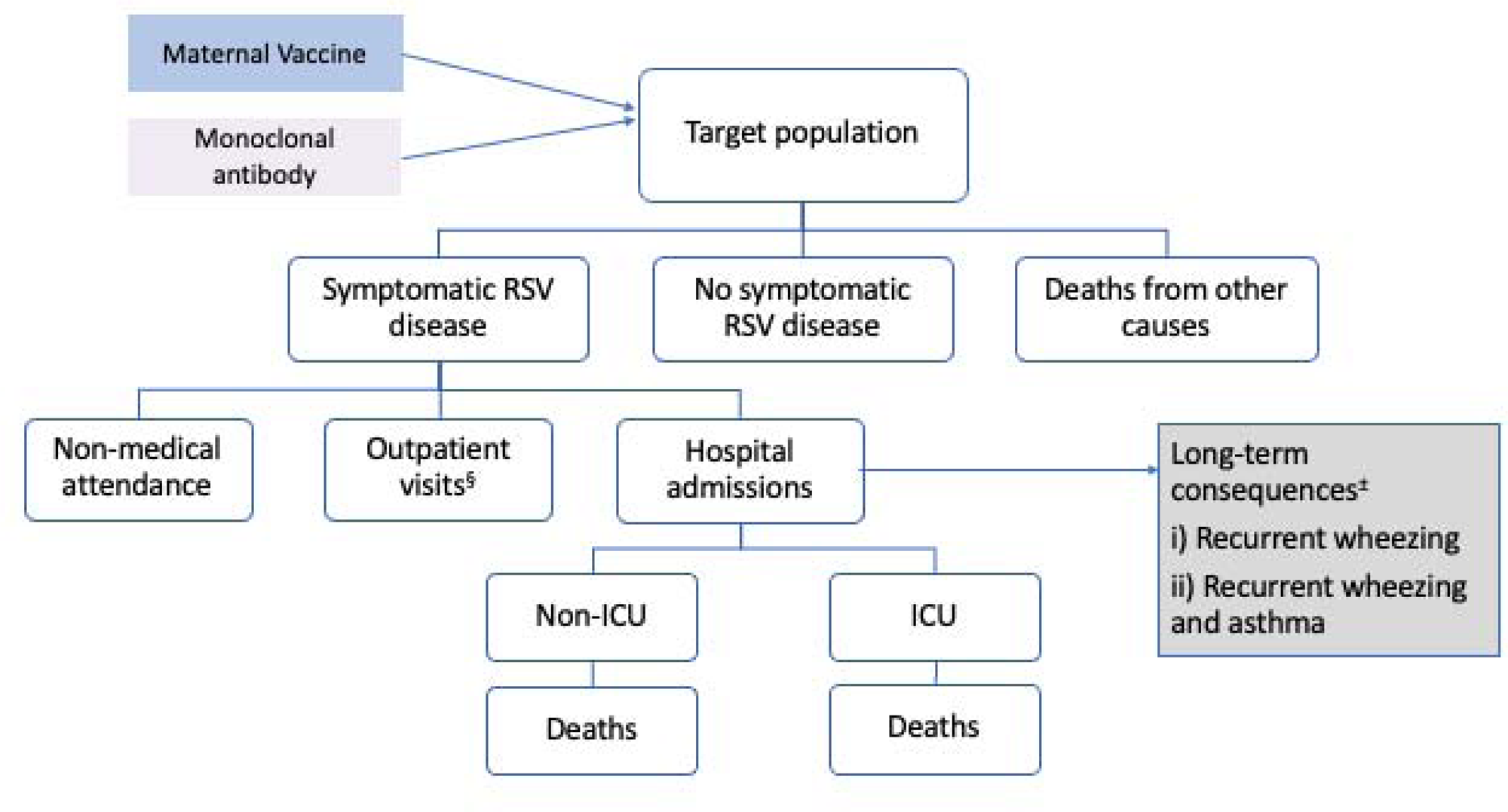
Model Structure. RSV: respiratory syncytial virus. § Outpatient visits include primary care consultations (either general practitioners or paediatrician consultation), hospital outpatient consultations and emergency department visits without hospital admission. ± Scenario analysis only: patients with RSV-related hospital admission within the first year of life have age-specific probabilities of (i) recurrent wheezing events up to 3 years of age and (ii) recurrent wheezing and asthma events up to 13 years of age.

**Figure 2.**
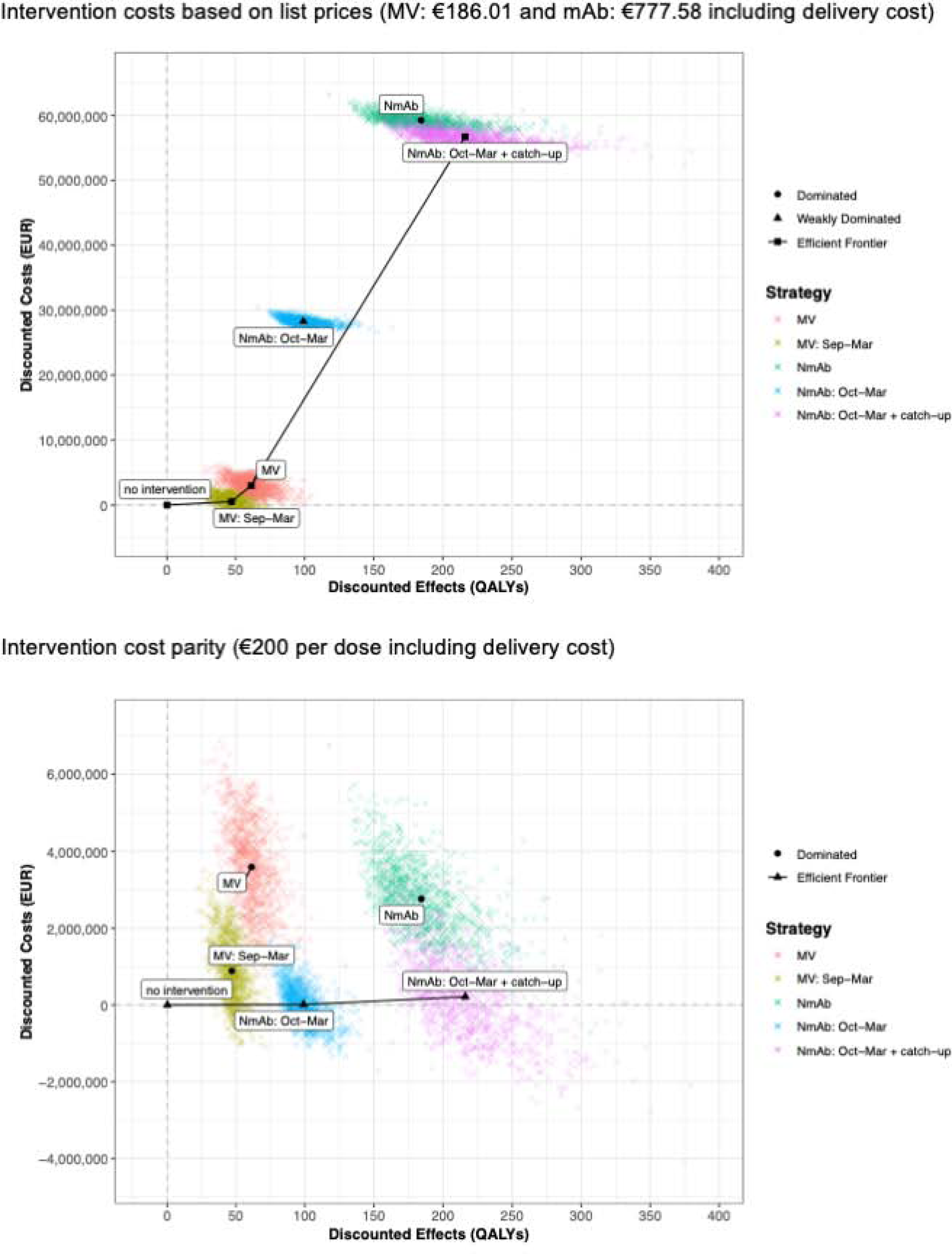
The cost-effectiveness plane from the HCP perspective. QALY: quality-adjusted life year, EUR: euro. HCP: health care payers’, MV: year-round maternal vaccine, MV: Sep-Mar: seasonal maternal vaccine from September to March, NmAb: year-round nirsevimab, NmAb: Oct-Mar: seasonal NmAb strategy from October to March, NmAb: Oct-Mar + catch-up: seasonal NmAb combined with a catch-up strategy.

Using list prices, the preferred strategy switched from no intervention to seasonal MV once the WTP ≥€11,276 per QALY gained (Figure S. 7). Under cost parity, the seasonal NmAb strategy was preferred at WTP <€96 per QALY gained, while the seasonal plus catch-up NmAb strategy became optimal if WTP ≥€1,725 per QALY (Figure S. 7). Detailed incremental cost-effectiveness ratios (ICERs) versus no intervention and the next best alternative are shown in Tables S.13 and S.14, respectively. Those findings were aligned with those in Figure 2, Table S.11 and Figure S. 7-8.

### Bivariate intervention cost analysis

All-inclusive per-dose intervention costs were varied, and the full incremental results, comparing 5 strategies to no interventions and to each other, are presented in Figure 3. From the HCP perspective, using an arbitrary WTP threshold of €35,000 per QALY gained, ‘no intervention’ (blue) was preferred when the costs of MV and NmAb exceeded €240 and €280 per dose, respectively. The seasonal plus catch-up NmAb strategy (yellow) became the preferred option if NmAb costs fell below €210, regardless of the costs of MV. When MV costed relatively less (<€75 per dose) and NmAb costed more (>€220 per dose), the year-round MV strategy (green) was optimal. If MV costed less than €230 and NmAb cost between €220 and €850, seasonal MV strategy (grey) became the preferred strategy.

**Figure 3.**
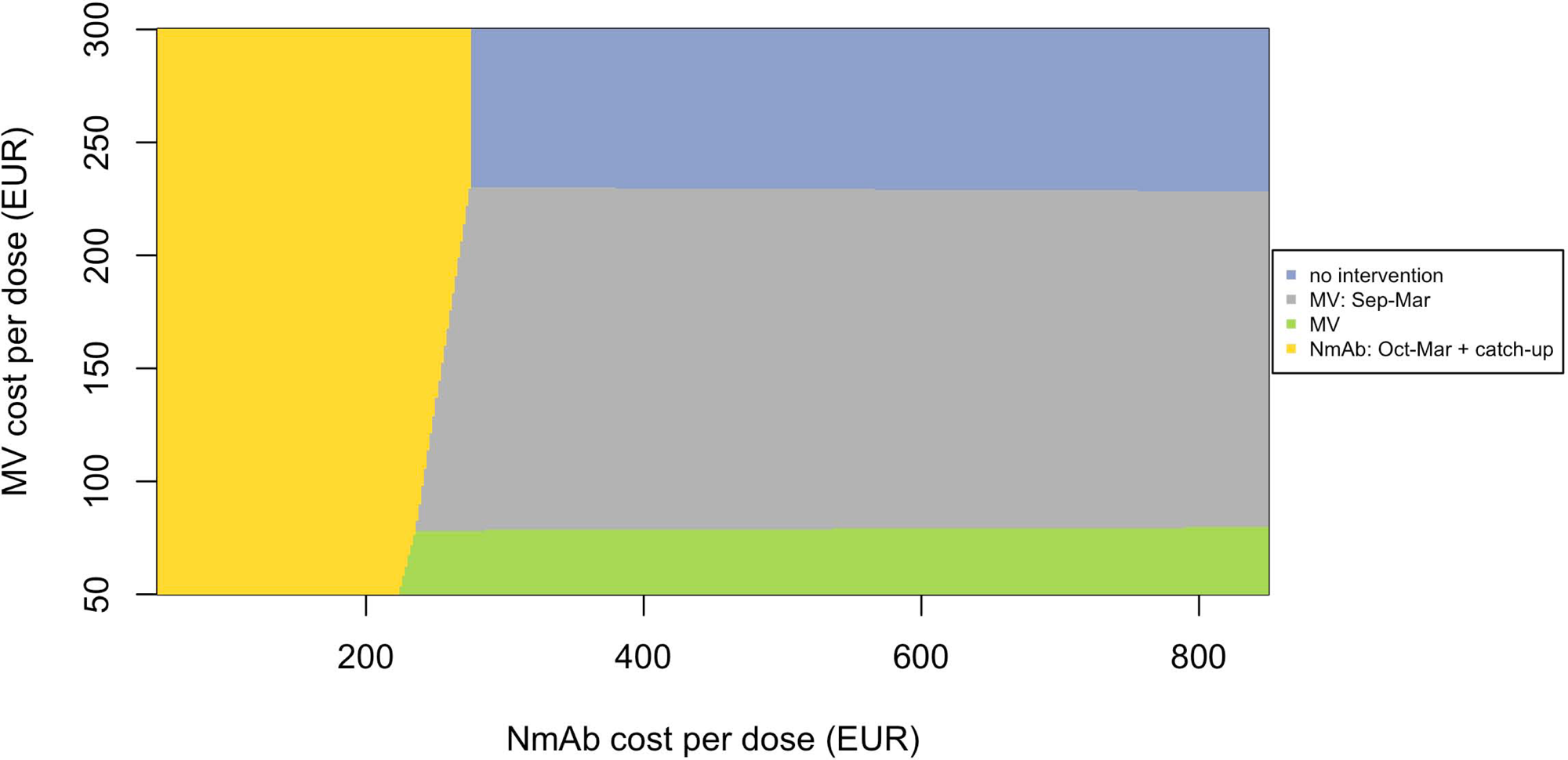
Intervention cost threshold analysis from the HCP perspective (all-inclusive cost per dose), comparing 5 strategies to no intervention and to each other, at willingness to pay of €35,000 per QALY gained. Y-axis: maternal vaccine costs ranged from €50-€30, X-axis: mAb costs from €50–€850. Each colour indicates a preferred RSV strategy, with the greatest uncertainty at the boundaries where strategies change. EUR: euro, HCP: health care payers’, QALY: quality-adjusted life-year, MV: year-round maternal vaccine, MV: Sept-Mar: seasonal maternal vaccine from September to March, NmAb: Oct-Mar: seasonal NmAb strategy from October to March, NmAb: Oct-Mar + catch-up: seasonal NmAb combined with a catch-up strategy.

Additional results for alternative thresholds of €0, €20,000, and €50,000 per QALY gained are provided in the Supplement (Figure S. 9). At a higher WTP threshold (i.e. €50,000), similar trends emerged, with the colour pattern shifting left and downward, allowing higher intervention costs to remain cost-effective. At a lower WTP threshold (i.e. €20,000), the pattern shifted right and upward, indicating lower intervention costs were needed for strategies to be preferred.

The seasonal NmAb strategy was not preferred, because the seasonal NmAb is less costly but also less effective than the seasonal NmAb plus catch-up strategy. At the WTP above €20,000 per QALY gained, the seasonal NmAb plus catch-up strategy was optimal due higher QALY gained. The seasonal NmAb strategy could be the preferred strategy only over a narrow mAb cost per dose range and given a WTP value of €0 per QALY gained. In other words, if policy makers are not willing to pay anything to gain QALYs in the population, then the seasonal mAb strategy without catch-up could be preferred, if the costs for NmAb are around €200 per dose and for MV >€160 per dose.

The year-round NmAb strategy was dominated by the seasonal plus catch-up NmAb strategy (Figure 2) and was therefore never preferred or shown in subsequent figures.

In scenario analysis, Figure 4 presents the inclusion of the ‘combined’ strategy alongside the other separate strategies at an arbitrary WTP threshold of €35,000 per QALY gained. The ‘combined’ strategy (pink) occupied the region where both NmAb and MV costs were relatively low, though assuming MV ≥€50 per dose. Given WTP of €35,000 per QALY, the ‘combined’ strategy was preferred when MV costed less than NmAb, with NmAb costing between €75 and €250 per dose. Additional results for thresholds of €0, €20,000, and €50,000 per QALY are in Figure S.10.

**Figure 4.**
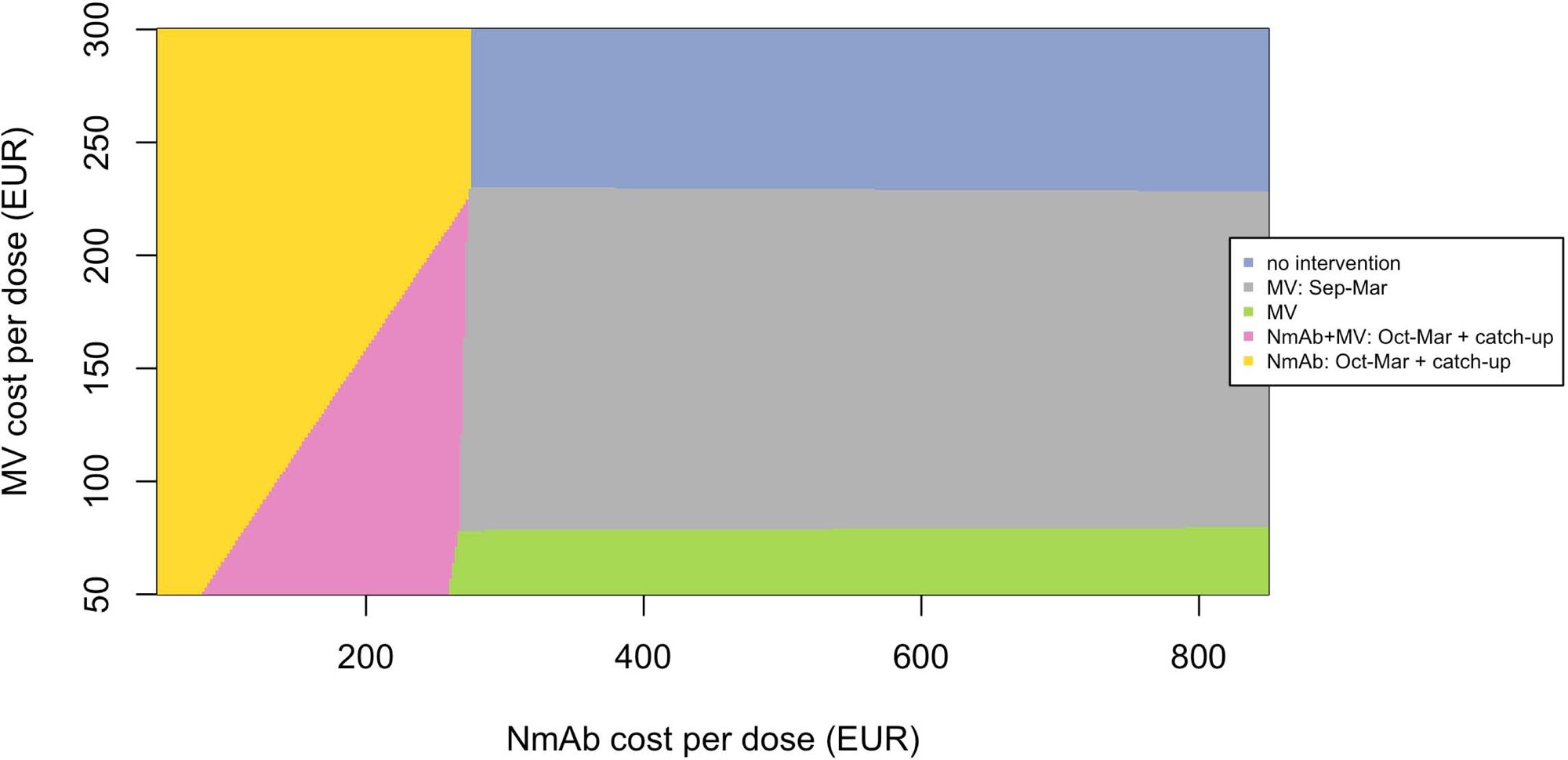
Scenario analysis: intervention cost threshold analysis from the HCP perspective (all-inclusive cost per dose), comparing 6 strategies including a ‘combined” strategy to no intervention and to each other, at willingness to pay of €35,000 per QALY gained. Y-axis: maternal vaccine costs ranged from €50-€30, X-axis: mAb costs from €50–€850. Each colour indicates a preferred RSV strategy, with the greatest uncertainty at the boundaries where strategies change. EUR: euro, HCP: health care payers’, QALY: quality-adjusted life-year, MV: year-round maternal vaccine, MV: Sept-Mar: seasonal maternal vaccine from September to March, NmAb: Oct-Mar: seasonal NmAb strategy from October to March, NmAb: Oct-Mar + catch-up: seasonal NmAb combined with a catch-up strategy, NmAb + MV: Oct-Mar + catch-up: seasonal MV supplemented with NmAb for infants born during the RSV season and of which the mother was not vaccinated, and an out of season NmAb catch-up programme.

Influential drivers identified by expected value of partial perfect information and scenario analyses

Expected value of partial perfect information (EVPPI) was estimated across WTP thresholds to identify key drivers of decision uncertainty (Figure S. 11). In the base case using list prices, the most influential parameters were efficacies of MV against hospitalisation and ICU admission, and age-specific outpatient incidence. Under cost parity, NmAb’s efficacy against hospitalisation and ICU (assumed equal based on pooled phase 3 RCTs) was the main driver, followed by outpatient incidence, and outpatient and hospitalisation costs. In both cases, uncertainties in efficacy against hospital and ICU admissions had a much greater impact than all other parameters.

A range of scenarios (Table S. 15) were analysed to explore sensitivity and uncertainty and identify key drivers. Compared to the HCP perspective, the societal perspective favoured the seasonal MV strategy at a lower WTP threshold using list prices and found the seasonal plus catch-up NmAb strategy to be cost-saving under the cost parity scenario (Figure S. 12).

As shown in Figure 5, including recurrent wheezing and asthma up to age 3 years and 13 years significantly affected the results. With list prices, the seasonal plus catch-up NmAb strategy became cost-effective around €22,000 (age 3) and €10,000 (age 13) per QALY. Under cost parity, it was cost-saving in both scenarios.

**Figure 5.**
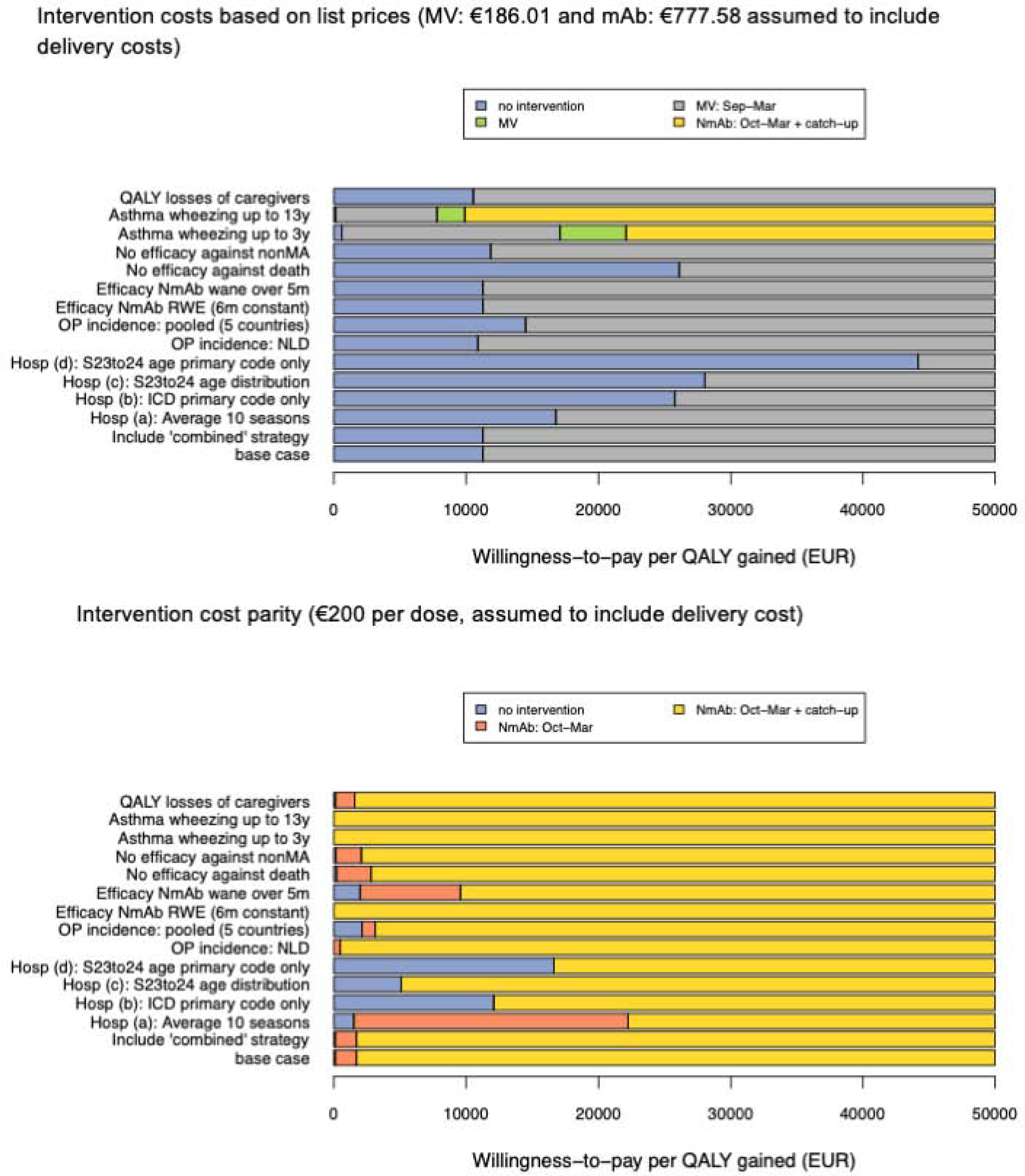
Deviation of scenario analyses from base case on the choice of preferred strategy by willingness to pay per QALY (from the HCP perspective). EUR: euro, HCP: health care payers, QALY: quality-adjusted life-year, MV: Sept-Mar: seasonal maternal vaccine from September to March, NmAb: Oct-Mar: seasonal NmAb strategy from October to March, NmAb: Oct-Mar + catch-up: seasonal NmAb combined with a catch-up strategy. Hosp: hospitalisation, QALY: quality adjusted life-year, y: year, m: month, ICD: international classification of diseases, NmAb: nirsevimab, non-MA: non-medically attended, RWE: real-world evidence, OP: outpatient, S: season, NLD: the Netherlands. S23to24: RSV season 2023/2024.

Alternative hospital burden scenarios (Figure 5) would lead to different findings. Hospitalisation burden in infants was estimated using four alternative approaches: (a) 10-season average, (b) primary diagnosis codes only, (c) 2022/2023 age distribution for infants aged 1–11 months, and (d) a combination of (b) and (c). Compared to the base case, the cost-effectiveness bars shifted progressively rightward from (a) to (d) approach, indicating that higher WTP thresholds were needed for the seasonal MV (list prices) and NmAb strategies (cost parity) to remain optimal.

Assuming no efficacy against RSV-related deaths led to large changes under list prices, with the seasonal MV strategy requiring a higher WTP (∼€27,000 per QALY) to be cost-effective. Under cost parity, the seasonal plus catch-up NmAb strategy needed only slightly higher WTP thresholds (∼€200 and ∼€2,000, respectively shown in Figure 5 bottom panel). In the base case using list prices, variations in NmAb efficacy had no effect at WTP values below €50,000 per QALY, as no NmAb strategy was cost-effective due to high list price. However, under cost parity (€200), applying pooled RWE effectiveness estimates (i.e. higher than the base case) with 6 months protection ^14^, made NmAb strategies cost-saving. When a shorter 5-month duration of protection was assumed ^15^, the seasonal NmAb strategy became cost-effective above €1,200 per QALY, while the seasonal plus catch-up strategy required a higher WTP to be preferred.

Other scenarios, including the use of Dutch and pooled outpatient incidence rates (vs. Spanish data in the base case), no efficacy against non-MA episodes, and inclusion of parental QALY losses, had limited impact on the overall results.

Figure 5 indicates that the ‘combined’ strategy had no impact on the cost-effectiveness results, whether using the list price or cost parity. NmAb’s list price was too high for either separate or combined strategies to be cost-effective. At cost parity, seasonal NmAb with catch-up, was preferred given its higher effectiveness compared with MV. This result was consistent with Figure 4, showing that when NmAb was too costly (>€250) or when MV cost the same as NmAb, the combined strategy was not preferred.

### Budget impact analysis

At 40% coverage, year-round and seasonal MV strategies were estimated to avert €5.1 million and €4.2 million in treatment costs, respectively (Table S. 16). Assuming a €50–€250 per-dose cost range, MV costs range from €2.1–€10.9 million (year-round) and €1.3–€6.3 million (seasonal). At 90% coverage, the seasonal NmAb and seasonal plus catch-up NmAb strategies were projected to reduce treatment costs by €9.8 million and €19.4 million, respectively (Table S.17). With per-dose costs ranging from €50 to €850, immunisation costs reached €2.4–€41.6 million for seasonal NmAb and €4.9–€83.1 million for the seasonal plus catch-up NmAb strategy.

The Return on Investment (RoI) depended on strategy and assumed cost per dose. Intervention options that use relatively more doses (i.e., year-round and catch-up programmes) require higher investments upfront (Table S. 16-17). As such, investment cost greater than prevented medical costs were expected unless all-inclusive per-dose costs fall below €110 for year-round MV, €160 for seasonal MV,

€200 for seasonal NmAb, and €190 for seasonal plus catch-up NmAb.

## Discussion

This analysis assessed the RSV burden in Belgian children under 5 years of age and evaluated the cost-effectiveness and budget impact of MV and NmAb protecting infants up to 6 months. Before the introduction of NmAb in the 2024–25 season, we estimated that RSV caused annually on average 8,638 non-ICU hospitalisations, 428 ICU admissions, 5 deaths, and 968 QALYs loss and €43 million in direct healthcare costs (without discounting). Overall, NmAb strategies are expected to prevent a greater RSV disease burden than MV strategies. This is not only due to the expectation that coverage of MV would be markedly lower than coverage of NmAb in the respective target groups (40% versus 90%, respectively), but also due to NmAb’s higher overall efficacy and effectiveness over time. Even when assuming equal coverage, NmAb remained more effective. However, with current list prices (NmAb: €777.58; MV: €186.01) ^12,13,16^, the increased effectiveness does not close the gap in terms of cost-effectiveness.

In the 2024–2025 season, NmAb was administered in hospitals to infants born during the RSV season via an age-based online application system ^17^. However, for those born outside the season, immunisation had not yet been implemented in Mother & Child Clinics (K&G and ONE). Our analysis showed that the seasonal plus catch-up NmAb strategy could achieve the greatest reduction in disease burden, but the out-of-season catch-up presents significant implementation challenges.

Administering it just before the RSV season would concentrate operational demands in September and October, covering nearly half the birth cohort. Successful implementation will require coordinated efforts, dedicated funding, and adequate resources. For MV, experience with maternal pertussis and influenza vaccination in Flanders indicates that gynaecologists typically refer patients to GPs via e-prescription rather than administer vaccines themselves. This referral process requires additional visits to both the pharmacy and GP, adding specific administration costs. The MV also has a narrow immunisation window (28–36 weeks gestation) and, while it can be given with seasonal influenza, it cannot currently be co-administered with the pertussis vaccine ^10^. A coordinated communication strategy may help improve overall coverage of seasonal maternal vaccines. Alternatively, the ‘combined’ strategy appears interesting in terms of cost-effectiveness, offering pregnant women/parents the choice of maternal vaccination, with the option for NmAb administration in cases of missed opportunities or other concerns. However, this approach would increase programme complexity and require healthcare providers to deliver clear, comprehensive information to support informed decision-making for individuals. Moreover, establishing a comprehensive RSV immunisation administration database is essential to prevent unnecessary repeated injections.

Beyond intervention costs, results were highly sensitive to RSV hospitalisation burden. The RSV hospitalisation rates in Belgian infants are among the highest in Europe ^2^ and more than double the global average for high-income countries ^18^. This may be partly caused by differences in tertiary care accessibility and patient management practices. Moreover, the number of hospital admissions increased over time prior to the COVID-19 pandemic, despite a gradual decline in the birth cohort size. This trend in reported numbers is likely attributable to several factors, including enhanced diagnostic testing and improved medical coding practices. Additionally, the implementation of other successful childhood (e.g., pneumococcal conjugate and rotavirus vaccination) and maternal (e.g., pertussis and influenza vaccination) immunisation programmes may have contributed to relatively greater availability of paediatric hospital beds during winter seasons, thereby facilitating more admissions.

The impact of RSV immunisation on wheezing and asthma remains uncertain, both in causality and measurement ^2,19–21^. However, when modelling potential long-term effects by linking severe infant RSV (proxied by hospitalisations) to later wheezing and asthma, the results showed a substantial influence, making the seasonal mAb plus catch-up strategy potentially cost-effective even at prices closer to the current list level.

Although an increasing number of RSV cost-effectiveness analyses have been published in Europe, most have focused on comparing RSV intervention(s) to standard care, with several estimating cost-effective price thresholds based on specific official or arbitrary WTP thresholds. Conducting economic evaluation without all relevant options can mislead policy. To address this, we conducted a full incremental analysis of all feasible strategies in Belgium, in consultation with national experts. As shown in Table S. 13 and S.14, the ICERs for the seasonal plus catch-up NmAb strategy were substantially lower (i.e. more attractive) when compared simply to no intervention rather than to the appropriate next best alternative strategy.

Our analysis can be compared with three previously published full incremental cost-effectiveness analyses: in Norway ^22^, in six European countries ^23^, in England ^24^. Our analysis found for comparable intervention options higher threshold intervention costs per dose at which these interventions can be considered cost-effective than Li et al.^22,23^ found for any of the seven countries using a similar static model structure in 2022 to 2023. The primary reason is that Belgium has substantially higher RSV-coded hospitalisation rates compared to these countries. Moreover, in our baseline analysis, in addition to assuming that these interventions prevent RSV attributable infant mortality, we were able to use more recent and more favourable information on efficacy and duration of protection (e.g., 6 months duration of protection for both NmAb and MV). Hodgson et al.^24^ used a dynamic transmission model, comparing four strategies both to ‘no intervention’ and to one another. They also performed a similar two-way threshold analysis using the UK’s official WTP threshold of £20,000 per QALY gained. They concluded that a seasonal NmAb programme could be cost-effective up to £84, while seasonal MV could be cost-effective up to £80 at all-inclusive cost. The main driver of the difference with our findings for Belgium was again the higher hospitalisation incidence rate used in our analysis. Moreover, the National Health Service costs per GP visit (£35), non-ICU admission (£1,100), and ICU admission (£2,905) were substantially lower than the corresponding costs in Belgium. Furthermore, Hodgson et al. attributed lower QALY losses for both MA and non-MA episodes ^24^.

This study offers several key strengths. It applied age-specific national disease burden data, representative cost and HRQoL inputs, and pooled clinical evidence from a systematic review to inform the model, supporting evidence-based decision-making. We incorporated a broad range of WTP thresholds and conducted advanced analyses probabilistic sensitivity (including value of perfect information and net loss analyses), scenario analyses, and bivariate price threshold analyses in order to enhance the relevance for policymakers, especially in price negotiations and public tender processes. We also used the latest real-world effectiveness data. Finally, we included a ‘combined’ strategy and identified the price ranges for both interventions under which it would become the preferred option.

While the disease burden may be underestimated in some respects, and thus the interventions’ impact somewhat understated, we expect this to remain limited. More importantly, our conservative approach generally ensures that any adjustment for the following limitations would likely strengthen the cost-effectiveness findings rather than weaken them. First, we used a static model that does not capture herd immunity. However, this limitation has also likely minimal impact given that (i) no intervention targets infants over 6 months, (ii) both MV and NmAb offer short-term protection, and (iii) infants primarily act as a “sink” rather than drivers of RSV transmission. Furthermore, a formal model comparison using both static and dynamic frameworks for similar RSV interventions showed comparable outcomes when the same input parameters were applied ^25^. Second, some RSV-associated illness, such as otitis media ^26^, was not accounted for in our analysis due to lack of data, and therefore the overall health and economic benefits of RSV prevention strategies may be underestimated. Also, we did not account for RSV protection in pregnant women receiving MV, slightly underestimating its impact. However, this is likely minimal as RSV is typically mild in healthy adults. Potential changes to preterm birth risks with MV have not been explored but are also likely to be limited since there is no conclusive evidence to this effect yet^27^. Finally, while our analysis used the current palivizumab programme as the ‘no intervention’ comparator, we did not include potential cost offsets from replacing it with NmAb in high-risk children. This omission is unlikely to affect the preferred strategy, since ‘no intervention’ is already dominated, but it may impact the budget impact analysis.

Overall, our analysis found the seasonal NmAb with catch-up strategy to be the most effective and the seasonal MV the least effective. However, at current list prices (€186.01 and €777.58 for MV and NmAb, respectively), only the seasonal MV strategy was cost-effective at a WTP threshold below €50,000 per QALY. Bivariate price threshold analysis showed that substantial reductions in per-dose immunisation costs are needed for most strategies to be cost-effective. Apart from interventions’ cost, results were highly sensitive to assumptions about RSV disease burden, especially hospitalisation rate and intervention efficacy against severe outcomes.

Future research should also focus on assessing the delivery cost and clarifying the causal link between RSV and subsequent wheezing and asthma.

## Methods

This section summarises our methods and key inputs (Table 1). More details are available in the Supplement.

**Table 1.**
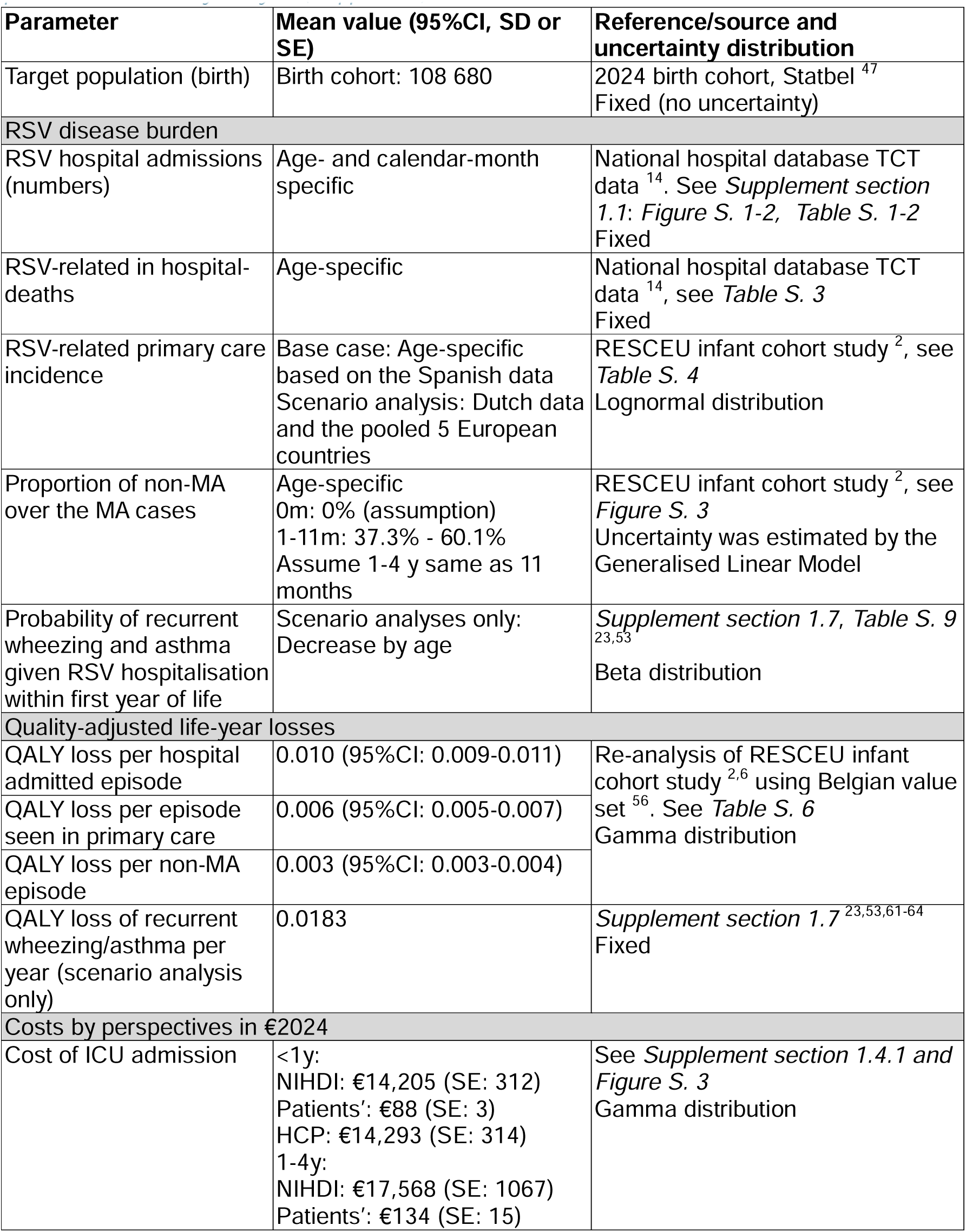
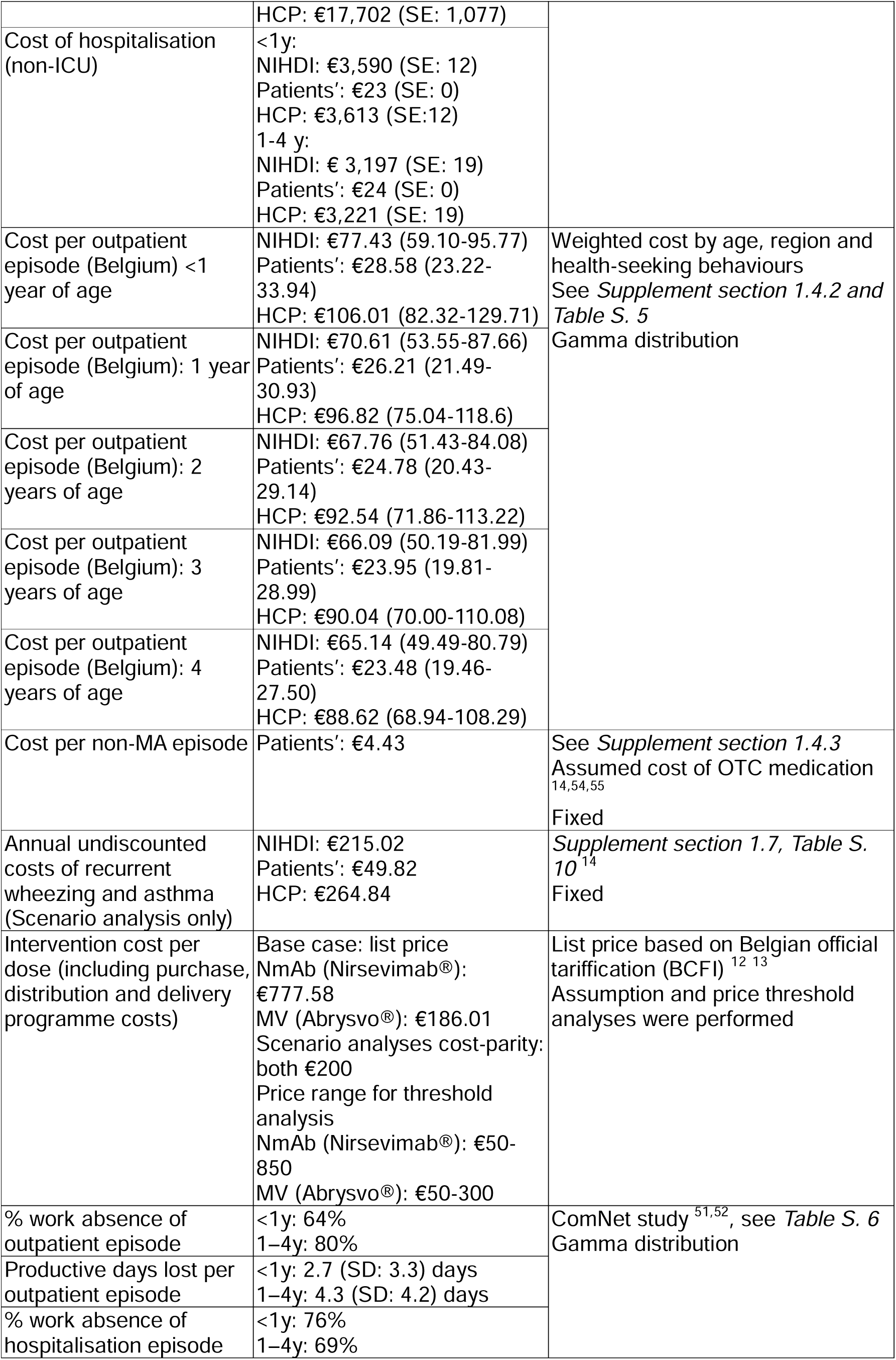
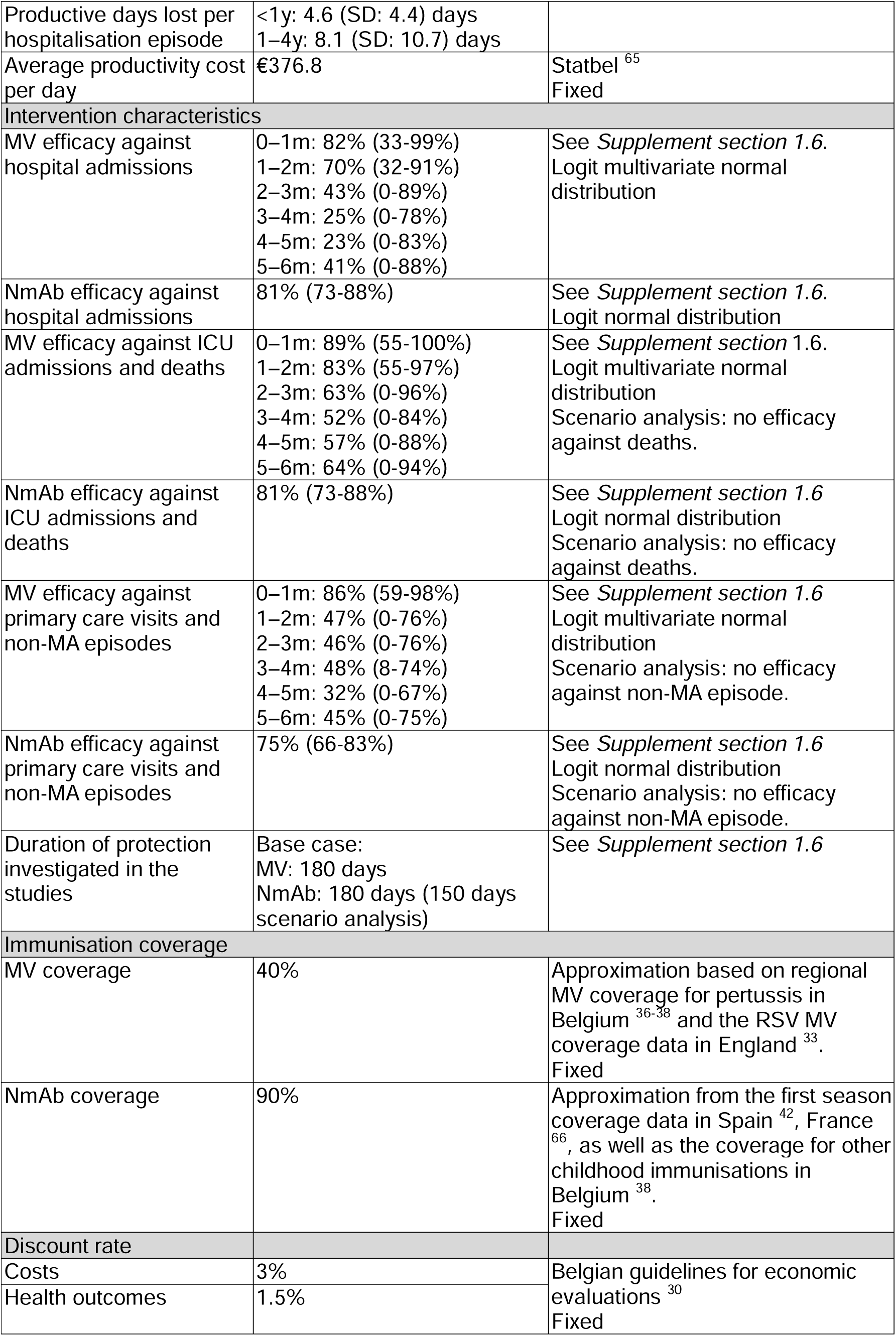
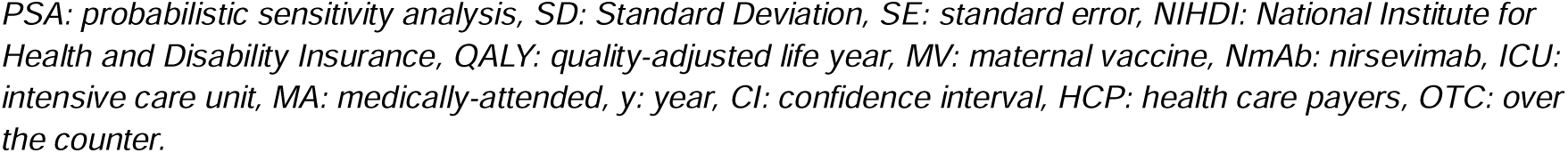
Input parameters used in the cost-effectiveness analysis and uncertainty distribution for the probabilistic sensitivity analysis (if applicable).

### Model structure

Static cohort model MCMARCEL ^22,23,28^ was adapted to the Belgian context to reach the objectives stated above. The chosen static modelling approach was in line with WHO guidance^29^, which considers to be appropriate when herd immunity effects are limited and positive, particularly if supported by evidence from model comparisons, as was the case here ^29^ ^25^. The Belgian birth cohort was followed monthly over a time horizon of 5 years, including both full-term and preterm infants (see Figure 1). The model incorporates QALY loss and healthcare costs linked to symptomatic RSV, including outpatient episodes, hospitalisations (with and without ICU), and non-medically attended cases. The evaluation followed Belgian guidelines for economic evaluation and budget impact analysis ^30,31^, adopting the health care payer perspective (federal government, federated authorities, and patients) in the base case. Uncertainty was explored with value of information analyses ^32^. In the scenario adopting a societal perspective, we included parental productivity losses due to RSV illness in their children using the human capital approach. However, the productivity losses due to premature deaths were not included to avoid double counting. Costs and QALYs beyond one year were discounted at 3% and 1.5%, respectively. All costs were adjusted to 2024 euro values using the healthcare consumer price index. In the absence of an explicitly defined threshold for the willingness to pay (WTP) for a QALY gain, we explored a range of WTP values (€0 to €50,000 per QALY).

### Immunisation strategies

We assumed 40% coverage for MV, informed by the first year of the seasonal RSV MV programme in England ^33^ and historical uptake for seasonal influenza and pertussis maternal vaccination in Belgium ^34–37^. For NmAb, a higher coverage of 90% was assumed, based on uptake levels of comparable early-life immunisations such as the hexavalent vaccine in Belgium ^38–41^, as well as the high NmAb uptake rates reported in Spain and France ^42–44^. Given all costs are included as variable costs in our static model-based analysis, coverage levels do not impact any of our cost-effectiveness estimates, but they do affect effectiveness and budget-impact estimates.

### Model input parameters and assumptions

We systematically searched PubMed to identify recent evidence on RSV burden, healthcare use, costs, and HRQoL in Belgium (details in Table 1 and ^14^). As highlighted in previous research ^3,5,45,46^, data on RSV in outpatient and community settings remain scarce due to limited RSV testing. We therefore included evidence from multi-country European studies and neighbouring countries, after discussions on contextual appropriateness with a Belgian expert panel (see acknowledgements). The most recent Belgian demographic data, including birth cohort size and age-specific ^47^ all-cause mortality rates ^48^, were sourced from the national statistical office (Statbel).

### RSV-ICD-coded hospitalisations

RSV-related ICU and non-ICU hospitalisations were analysed using Belgium’s national Technical Cell – Cellule Technique (TCT) database. The TCT obtains anonymised data from all Belgian hospitals and links it with sickness fund billing records. This analysis identified 118,025 episodes in children under 5 years over 14 calendar years (2008–2014 and 2016–2022, 2015 was excluded due to fundamental data issues) ^14^. Overall, RSV-related hospitalisations show clear seasonality from October to March, except during the COVID-19 pandemic, when an atypical peak occurred in May during 2020/21 and hospitalisation numbers remained low without a clear peak in 2021/22.

Over 10 complete pre-COVID-19 pandemic RSV seasons (2008/09 to 2013/14 and 2016/17 to 2019/20), an average of 8,300 RSV-coded hospital admissions occurred annually, with a rising trend despite a decreasing birth cohort. The most recent 4 pre-COVID-19 seasons (2016/17–2019/20), which used consistent ICD-10 coding, were deemed most representative for the post-pandemic period by the Belgian experts and used for our base case (Figure S. 1 and Table S. 1). Because the TCT database aggregates children aged 1–11 months, we used BELSARI-NET data, a 10-hospital surveillance network, which provided age-specific RSV laboratory confirmed cases from 2023–2025. Data from the 2018/2019 season were used to estimate monthly age-specific RSV hospitalisations (Table S. 2 and Figure S.2) ^49^. Various scenarios of using this information were tested.

### RSV-ICD-coded deaths in hospital

The RSV-coded in-hospital case fatality rate (hCFR) was estimated using 14 years of TCT data, with 60 RSV-related deaths recorded among Belgian children under 5 years, 27% of which occurred outside the ICU. All RSV-related deaths were assumed to occur in hospital, and age-specific hCFRs (Table S. 3) were applied. Since there is disagreement among clinical experts about the inevitability of infant mortality through RSV immunisation, we also performed scenario analysis assuming no deaths would be preventable by RSV immunisation.

### Non-hospitalised RSV

No age-specific incidence rates for RSV-related outpatient consultations, including primary care or emergency department visits, were identified through a published systematic review ^50^, and our own supplementary review. The RESCEU infant study provided both medically attended (MA) RSV-ARI and hospitalisation rates, allowing us to estimate the outpatient rate per country by subtracting the latter from the former, using a lognormal sampling approach (Table S. 4) ^2^. The ComNet study followed children over 3 RSV seasons (2020–2023) in 5 European countries including Belgium ^51,52^. Given similarities in healthcare use as reported by ComNet 1 to 4-year-olds matched that of the 6–11-month age group.

We also used RESCEU infant study data to estimate age-specific non-MA RSV symptomatic cases as a proportion of all medically attended cases in children under 1 year ^2,6^ (Figure S. 3).

### Resource use, direct and indirect costs

Hospitalisation costs were estimated using TCT data over 14 calendar years, with average costs stratified by age and ICU status. No major cost differences were observed before and after the COVID-19 pandemic (Figure S. 4). Out of the average total cost of €14,293 per ICU and €3,613 per non-ICU hospital episode in infants (<1 year), most of the costs were covered by national health insurance, while patients co-paid an average of €88 and €23 per ICU and non-ICU hospital episode.

In Belgium, both general practitioners (GP) and paediatricians are directly accessible, so we accounted for the age-and region-specific distribution of consultations between these primary care providers, applying the corresponding unit costs per visit. Age-specific data on the number of visits per RSV outpatient episode and associated medication costs were obtained from the ComNet study ^51^. The estimated outpatient costs per MA and non-MA episode are shown in supplement (Table S.5). For non-MA cases, over-the-counter medication costs of €4.43 were included ^53–55^.

Productivity losses, based on Belgian self-reported parental work absence data from ComNet ^51^, were applied only to RSV cases >3 months, assuming no such losses during the initial 3 months, given typical maternity leave periods in Belgium (Table S.6).

### Health-related quality of life loss

QALY losses due to RSV infection, based on the RESCEU infant study ^6^, were calculated using the Belgian EQ-5D value set (Table S.7). As per the Belgian guideline, our base case included only infant QALY losses, but parental losses were also added in scenario analysis. Age-specific health utility values from the population norm ^56^ were applied to the life-years lost due to RSV mortality, to obtain QALY losses due to RSV-related deaths.

### Interventions’ characteristics: efficacy, durability and delivery costs

A bespoke systematic review of the literature on efficacy, effectiveness and safety of NmAb and MV was made and can be consulted elsewhere ^14^. The efficacy of NmAb was assessed at 150 days in phase 2b/3 trials ^57–59^, but the phase 3b trial demonstrated 180-day efficacy of NmAb against hospitalisation ^60^. The efficacy values of MV were reported at monthly intervals up to 180 days ^60^. Based on expert consensus, the latest trial data were used, in the base case assuming constant 6-month protection for NmAb and time-varying monthly efficacy for MV (Table 1 and Figure S. 5-6). These assumptions were subjected to sensitivity analysis (see below and Supplement section 1.6.2).

Distribution and delivery costs, including administration, storage, cold chain, awareness campaigns and wastage, for both products across Belgian regions could not be estimated due to regionally different uncertainties in how the RSV programme of choice would be integrated in the existing programmes. Therefore, we used an all-inclusive per-dose cost approach, through which all purchase and delivery associated costs would be included. Since the negotiated Belgian price of the RSV products is unknown for non-high-risk infants, but expected to be lower than the public list price, we used the current list prices (€186.01 and €777.58 for MV and NmAb, respectively) as a benchmark in the baseline, assuming they include all implementation and delivery costs, and explored also a cost-parity scenario assuming €200 per dose (all-inclusive cost).

### Probabilistic sensitivity and scenario analyses

Uncertainty in our economic evaluation was included via a probabilistic sensitivity analysis (PSA) with 1,000 samples to obtain cost-effectiveness acceptability curves (CEACs), and the expected value of perfect information (EVPI) over a range of WTP values. The EVPPI was used to identify key drivers and quantify the value of reducing their uncertainty.

Extensive scenario analyses were conducted to account for non-parameterised uncertainty, using alternative data selections and/or sources in relation to hospital burden, efficacy, waning, and durability (Table S. 8), as well as adopting a broader societal perspective and including parental QALY losses (Supplement section 1.6.2). We also assessed the impact of recurrent wheezing (up to age 3) and wheezing with asthma (up to age 13) in infants hospitalised for RSV in their first year, in line with Li et al. (2022) ^22^, using the relevant QALY and cost estimates from Belgium (Supplement section 1.7).

### Budget impact analysis

A budget impact analysis was conducted in accordance with Belgian guidelines to assess the affordability of implementing a new RSV strategy. Moreover, the return on investment (RoI) ratios were calculated by comparing net savings from the intervention to its cost.

## Declarations

### Data availability

All data generated or analysed during this study are included in this published article and its supplementary files, and the KCE report 402 ^14^.

### Code availability

The underlying code for this study is not publicly available but may be made available to qualified researchers on reasonable request from the corresponding author (XL) or the senior author (PB).

## Acknowledgements

The authors would like to thank Nivel: Valerie Sankatsing and Jojanneke van Summeren (Nivel) for providing additional analysis of the ComNet study; Hanmeng Xu and Daniel Weinberger (Yale University) for providing mean of the estimated effectiveness for the nirsevimab in the test-negative case-control study, Louis Bont (Wilhelmina Children’s Hospital in the University Medical Center Utrecht) for granting permission to re-estimate the quality-of-life data collected from the RESCEU prospective birth cohort study.

The authors also like to thank Marc De Falleur (INAMI – RIZIV); Yinthe Dockx (Sciensano); Pierre Hubin (Sciensano); Marc Raes (Belgian Academy of Paediatrics) and BELSARI-NET research group https://www.sciensano.be/fr/file/belsari-net-research-group (Sciensano) for providing data and expert opinions, and Olivier Ethgen (Université de Liège (Uliège); Maarten Postma (Rijksuniversiteit Groningen); Daan Van Brusselen (Ziekenhuis aan de Stroom (ZAS)) for their critical review of the KCE study report and valuable comments.

## Authors’ contribution

PB and DR initiated and supervised the study. XL, LW, DR, JB and PB conceptualised the study. XL analysed input data, performed a literature review and auxiliary data collection. ZM performed the literature review as the second reviewer, collected and analysed the health-related quality of life data. LW, XL and JB wrote the cost-effectiveness model codes. XL performed the cost-effectiveness analyses. CDM conducted national database analyses. NT collected and validated data in Belgium. DCZ conducted the systematic review and meta-analysis on efficacy, effectiveness and safety of RSV intervention. PB, LW, DR, NT, CDM, DCZ advised on model parameters, intervention characteristics and scenario analyses. XL, LW, PB wrote the initial manuscript draft. All authors critically reviewed the manuscript and provided final approval of the manuscript.

