## Supplement for "Cost-effectiveness of maternal vaccine and/or monoclonal antibody strategies against respiratory syncytial virus in Belgian infants"

Affiliations:

Address:

Campus Drie Eiken, Universiteitsplein 1

2610 Antwerpen, Belgium

### 1 Table of Contents

***1. Supplement Methods*** ***3***

**1.1 RSV-ICD-coded hospitalisations** **3**

**1.2 RSV-ICD-coded death in hospital** **6**

**1.3 RSV-related non-hospitalised consultations and RSV non-medically attended episodes** **7**

**1.4 Resource use, direct and indirect costs** **7**

1.4.1 Inpatient setting 8

1.4.2 Outpatient setting 8

1.4.3 Non-medically attended (non-MA) episodes 9

1.4.4 Cost associated with productivity losses 10

**1.5 Health-related quality-of-life** **10**

**1.6 Interventions' efficacy and durability** **10**

1.6.1 Base case: efficacy values based on clinical trials. 10

1.6.2 Scenario analysis: effectiveness based on real-world studies 12

**1.7 Long-term consequences** **13**

***2 Supplement results*** ***15***

**2.1 RSV-related disease and economic burden in Belgian children under 5 years** **15**

**2.2 Impact of interventions on RSV disease and economic burden** **15**

**2.3 Cost-effectiveness of RSV immunisation strategies** **16**

2.3.1 Cost-effectiveness of each strategy versus standard of care 16

2.3.2 Incremental cost-effectiveness plane: full incremental analysis 17

2.3.3 A full incremental analysis compared all five RSV strategies to no intervention and each other; ICERs excluding dominated strategies are reported in 18

2.3.4 Bivariant threshold analysis of interventions' cost 19

2.3.5 Expected value of partial perfect information 22

2.3.6 Scenario analyses 23

**2.4 Budget impact analysis** **25**

***3 Reference*** ***27***

### 1. Supplement Methods

A health economic analysis plan was developed at the project proposal and agreed at the start of the project. The plan guided all methodological and analytical decisions, although it is not publicly available. This analysis followed its predefined methods and assumptions, using the most up-to-date clinical, epidemiological, quality-of-life and economic data.

#### 1.1 RSV-ICD-coded hospitalisations

RSV-related hospitalisations coded with ICD codes were analysed using the Belgian national 'Technical Cell – Cellule Technique' (TCT) database, which links anonymised Minimal Hospital Data (MZG–RHM) to hospital reimbursement data from the Sickness Funds [1]. The total number of RSV-ICD-coded hospitalisations was assessed, with monthly and yearly counts shown in Figure S. 1. Episodes were identified using relevant primary and secondary ICD-9/10 codes from January 2008–December 2014 and January 2016–December 2022.

In the TCT dataset, children aged 1–11 months were grouped together. Age-specific monthly distribution of RSV hospitalisations was estimated using BELSARI-NET data on RSV-confirmed admissions, collected since 2012 from a network of 6 hospitals, expanded to 10 in 2023. Table S. 2 presents RSV hospitalisations in infants under 1 year for the 2018/2019 (pre-COVID-19) and 2023/2024 (peri-COVID-19) seasons, highlighting a notable shift in age distribution.

The 2018/2019 season was therefore used in the base case to redistribute non-ICU and ICU hospitalisations among infants aged 1–11 months (Figure S. 2). Scenario analysis used 2023/2024 (peri-COVID-19) data to assess the potential impact of an age shift in RSV hospitalisations.

Various scenarios of using this information were tested, given that the TCT data revealed an average annual RSV hospitalisation rate of 49.3 per 1,000 infants under 1 year, which was more than double the rate reported in the Global Burden of Disease study for high-income countries (22 per 1,000) and substantially higher than the 10.4–24.8 per 1,000 range observed in the REspiratory Syncytial virus Consortium in EUrope (RESCEU) infant study across 5 European countries [2]. For the 2023/2024 RSV season, a shift in age distribution among infants was observed based on BELSARI-NET, with fewer hospitalisations in the 0–2 month age group (40% vs. 50%), likely reflecting changes in social contact patterns during the COVID-19 period.

*Figure S. 1 – Number of RSV-ICD-coded hospitalisations in children <5 years by months from 2008-2022**

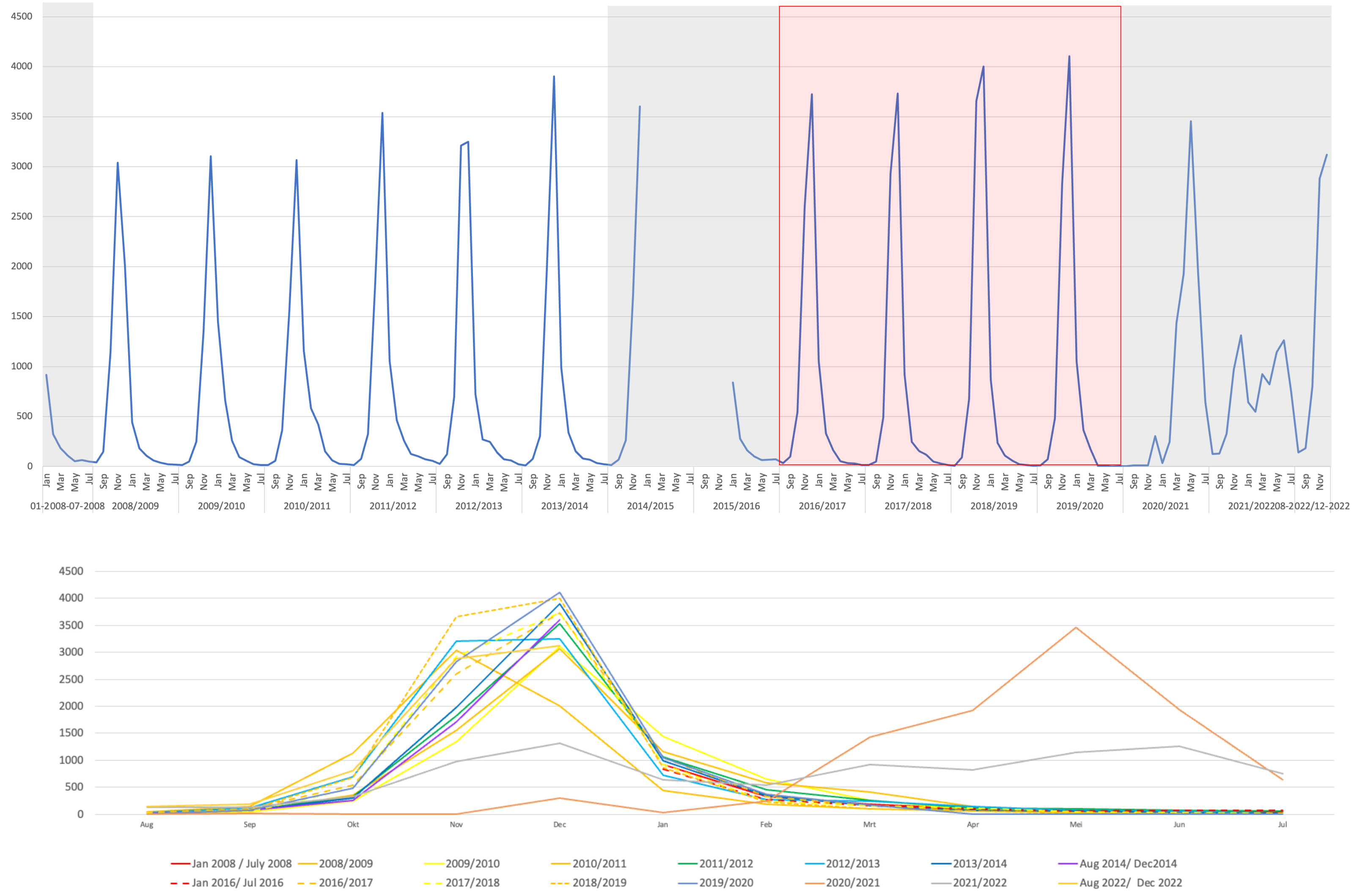


** TCT data is unavailable in calendar year 2015 due to technical errors. Seasons excluded from the analysis are shown as grey areas (top graph) and dashed lines (bottom graph). The seasons included as base case are shown as red shaded area (top graph).*

*Table S. 1 – Average number of RSV-ICD-coded non-ICU and ICU admissions in children <5 years by calendar month prior to the COVID-19 pandemic (average of 4 RSV seasons 2016/2017-2019/2020*)*

| Age | JAN | FEB | MAR | APR | MAY | JUN | JUL | AUG | SEP | OCT | NOV | DEC |
|---|---|---|---|---|---|---|---|---|---|---|---|---|
| Non-ICU hospital admissions | | | | | | | | | | | | |
| 0 months | 93 | 21 | 14 | 6 | 3 | 2 | 1 | 2 | 6 | 29 | 158 | 284 |
| 1-11 months | 657 | 193 | 97 | 37 | 16 | 11 | 6 | 8 | 50 | 332 | 1 815 | 2 491 |
| 1 year | 5 | 9 | 8 | 5 | 3 | 2 | 2 | 3 | 9 | 97 | 550 | 615 |
| 2 years | 118 | 35 | 17 | 6 | 3 | 2 | 1 | 1 | 5 | 38 | 216 | 207 |
| 3 years | 38 | 15 | 6 | 2 | 1 | 1 | 0 | 0 | 2 | 22 | 108 | 81 |
| 4 years | 11 | 5 | 2 | 1 | 1 | 0 | 0 | 0 | 2 | 7 | 28 | 27 |
| ICU admissions | | | | | | | | | | | | |
| 0 months | 16 | 7 | 3 | 1 | 0 | 0 | 0 | 1 | 3 | 7 | 34 | 60 |
| 1-11 months | 32 | 8 | 3 | 3 | 1 | 1 | 0 | 1 | 1 | 16 | 77 | 111 |
| 1 year | 1 | 0 | 0 | 0 | 0 | 0 | 0 | 1 | 0 | 1 | 8 | 11 |
| 2 years | 3 | 1 | 1 | 0 | 1 | 0 | 0 | 0 | 0 | 1 | 5 | 3 |
| 3 years | 1 | 0 | 0 | 0 | 0 | 0 | 0 | 0 | 1 | 0 | 3 | 1 |
| 4 years | 1 | 0 | 0 | 0 | 0 | 0 | 0 | 0 | 0 | 0 | 1 | 1 |

*Source: TCT data. * TCT data is unavailable in calendar year 2015 due to technical errors. ICU: intensive care unit.*

*Table S. 2 – Percentage of RSV hospitalisations per month since birth in children <1 year in the BELSARI-NET database*

| Age | 2018/2019 (pre-COVID-19) | 2023/2024 (peri-COVID-19) |
|---|---|---|
| 0m | 8.50% | 9.70% |
| 1m | 19.50% | 14.20% |
| 2m | 21.60% | 15.90% |
| 3m | 10.60% | 11.50% |
| 4m | 8.50% | 5.30% |
| 5m | 6.80% | 9.70% |
| 6m | 5.50% | 8.00% |
| 7m | 5.50% | 6.20% |
| 8m | 3.00% | 6.20% |
| 9m | 4.20% | 8.80% |
| 10m | 3.40% | 2.70% |
| 11m | 3.00% | 1.80% |

*m: month.*

*Figure S. 2 – Average number of RSV-ICD-coded hospitalisations (including both non-ICU and ICU admissions) by age and calendar month in children <5 years prior to the COVID-19 pandemic (2018/2019 season)*

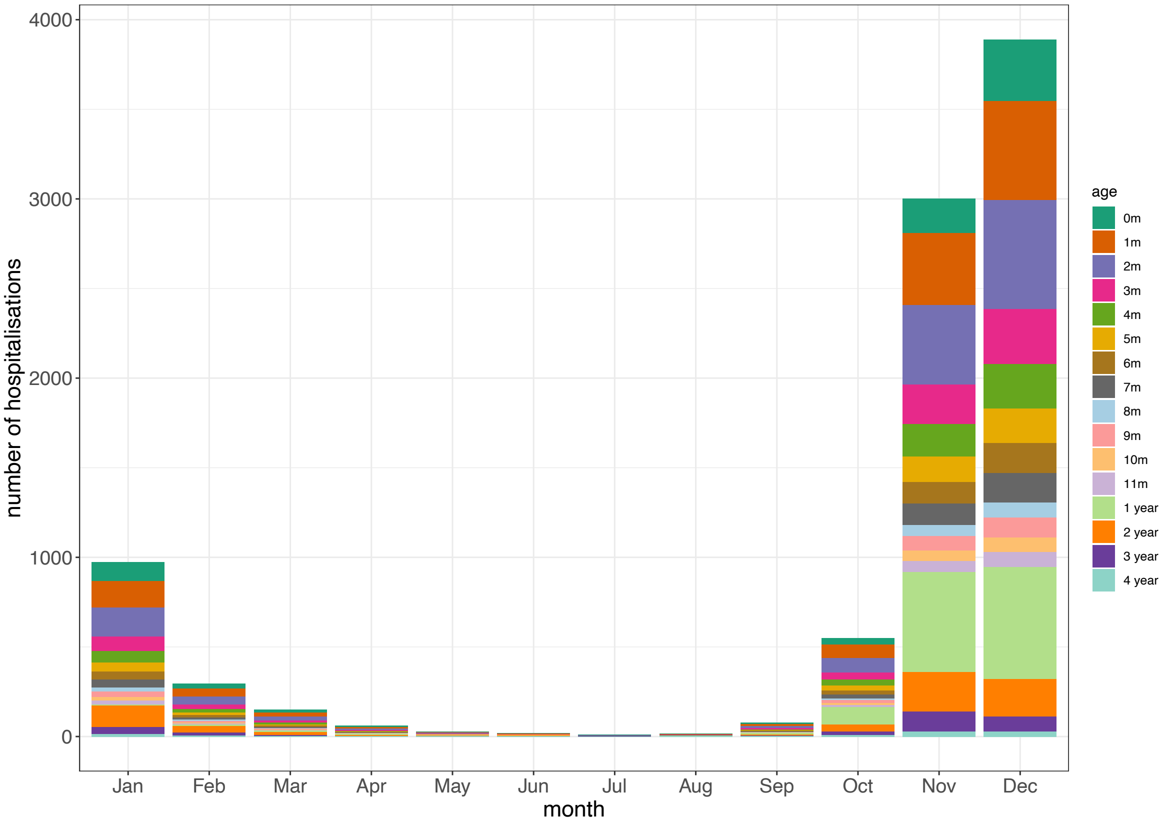


*m: month.*

#### 1.2 RSV-ICD-coded death in hospital

RSV-coded in-hospital case fatality rate (hCFR) was analysed using the TCT database over 14 years, with no distinct pre- or post-COVID-19 pattern observed [1]. Among Belgian children under five, 60 RSV-related deaths were recorded (Table S. 3), 27% of which occurred outside the ICU.

*Table S. 3 – Age-specific RSV-coded deaths and hCFR over 14 calendar years (2008-2014 and 2016-2022*)*

| Age | Hospitalisation without ICU admission | | | Hospitalisation with ICU admission | | |
|---|---|---|---|---|---|---|
| | **deaths** | **admissions** | **hCFR** | **deaths** | **admissions** | **hCFR** |
| <1m | 3 | 12 158 | 0.025% | 6 | 1 564 | 0.384% |
| 1-11m | 6 | 68 077 | 0.009% | 27 | 2 681 | 1.007% |
| 1y | 3 | 17 271 | 0.017% | 4 | 291 | 1.375% |
| 2y | 2 | 9 931 | 0.020% | 6 | 161 | 3.727% |
| 3y | 2 | 4 386 | 0.046% | 1 | 78 | 1.282% |
| 4y | 0 | 1 390 | 0.000% | 0 | 37 | 0.000% |
| Total | 16 | 113 213 | 0.014% | 44 | 4 812 | 0.914% |

** TCT data is unavailable in calendar year 2015 due to technical errors. hCFR: in-hospital case fatality ratio, ICU: intensive care unit, m: month, y: year.*

#### 1.3 RSV-related non-hospitalised consultations and RSV non-medically attended episodes

In the absence of Belgium-specific RSV outpatient incidence data, estimates were derived from the RESCEU infant study [2]. Full methodological details (i.e. assumptions, calculation, sampling) in the Belgian health care knowledge center (KCE) report [1].

Due to similar healthcare-seeking behaviour and paediatric care in Belgium and Spain, supported by data from ComNet study [3], we used the estimated RSV outpatient incidence rate from Spain (based on the RESCEU study) as the base case for Belgium (Table S. 4). Scenario analyses also considered age-specific outpatient rates from the Netherlands and pooled estimates from all five RESCEU countries [2].

*Table S. 4 – Age-specific RSV-associated outpatient ARI rate per 1,000 person-year (estimated mean and 95%CIs)*

| per 1,000 person-year | <3 month | 3–5 month | 6–11 month |
|---|---|---|---|
| Spain (Base case) | 205.87 (71.82-445.14) | 239.23 (93.44-488.44) | 108.17 (38.41-234.4) |
| The Netherlands | 237.73 (84.68-508.39) | 173.92 (52.44-415.72) | 245.17 (127.58-415.26) |
| Overall (5 countries) | 103.91 (52.81-174.69) | 165.29 (101.63-248.39) | 122.91 (82.95-172.24) |

Using non-medically attended (non-MA) episode data from the RESCEU infant cohort, we fitted a generalized linear model to estimate age-specific proportions of non-medically attended RSV cases [2,4], assuming there is non-MA episode in those under 1 month, and applying the 11-month rate to the 1–4-year group (Figure S. 3).

*Figure S. 3 – Proportion of non-MA RSV symptomatic cases over the MA symptomatic cases*

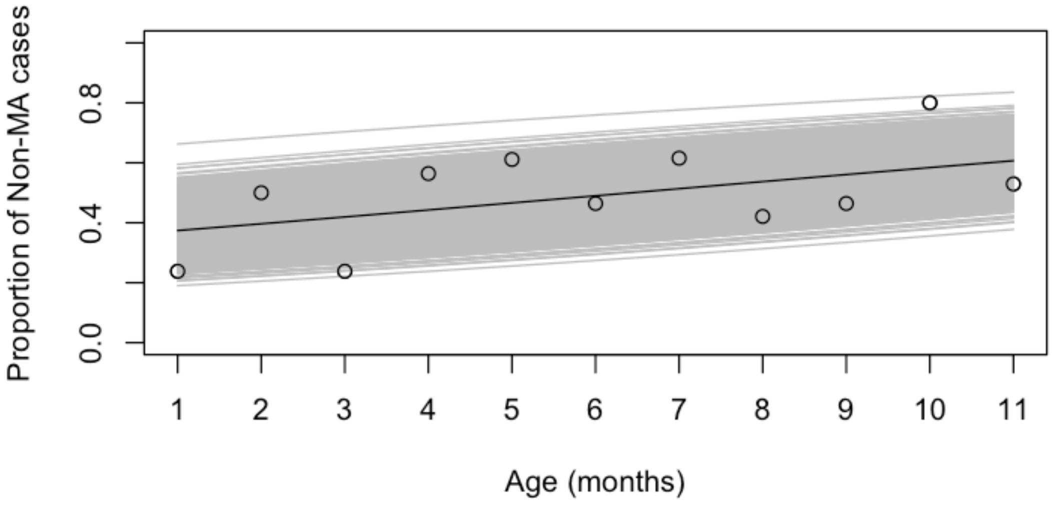


*Dots represent the data observed in the European observational study (RESCEU),[2,4,5] the grey lines represent the fitted values, and the solid black line represents the mean of the fitted value.*

#### 1.4 Resource use, direct and indirect costs

Following the Belgian Pharmacoeconomics guidelines, all costs were valued at the 2024 price level and reported from the perspectives of the National Institute for Health and Disability Insurance (NIHDI), patients, and healthcare payers (HCP) [6,7]. The uncertainty associated with the cost parameters was modelled using gamma distributions, where $\alpha = (\text{mean})^2/SD^2$ and $\beta = SD^2/\text{mean}$.

##### 1.4.1 Inpatient setting

Figure S. 4 shows the estimated the average cost per non-ICU and ICU admission over 14 years (appropriately inflated).

*Figure S. 4 – Average RSV-coded hospitalisation costs per admission and per year for ICU (top panels) and non-ICU admissions (bottom panels) in <1 year olds (left panels) and 1-4 year olds (right panels) (€ 2024)*

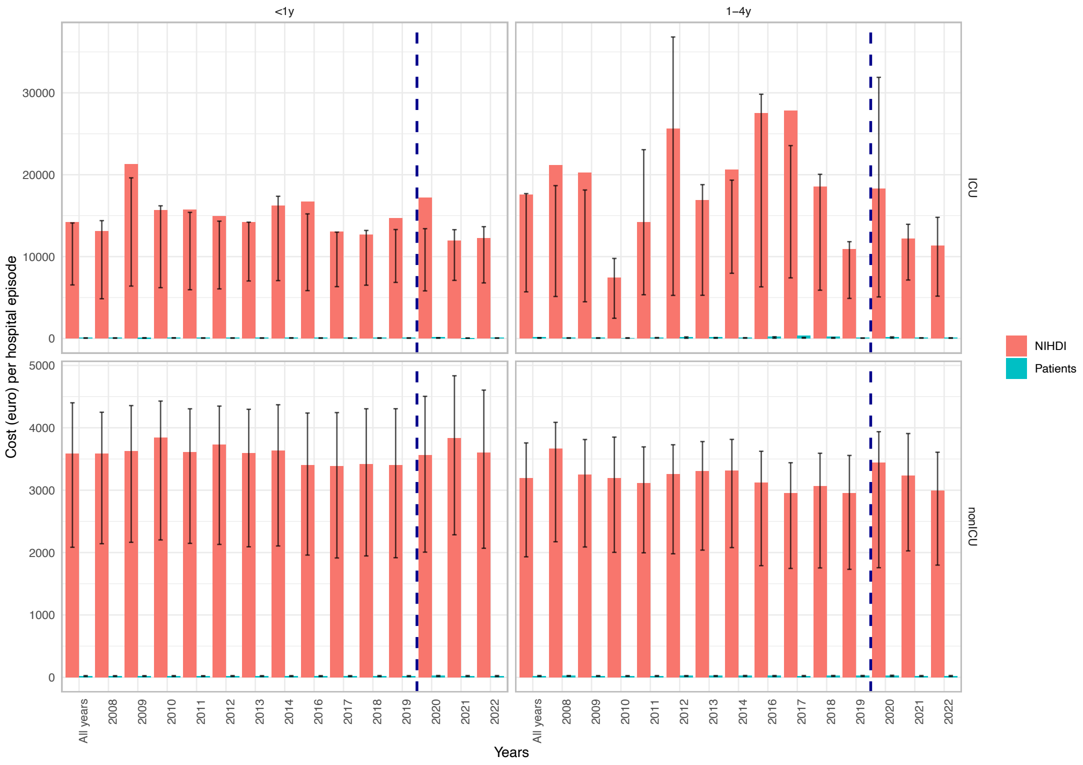


*The error bars present the 25th and 75th percentiles. The blue dotted line indicates before and after the COVID-19 pandemic. NIHDI: National Institute for Health and Disability Insurance.*

##### 1.4.2 Outpatient setting

The cost per RSV outpatient episode was calculated using Equation 1. The number of outpatient visits was from the ComNet study [3]. In Belgium, infants under 6 months had a mean of 2.8 visits (SD: 2.3), those aged 6–11 months had 2.5 (SD: 2.8), and 53.8% of children aged 1–4 years had multiple visits, with a mean of 2.3 (SD: 2.0) visits [3]. Unit costs for paediatricians (codes: 102071, 102572) and GPs (101032, 101076) were based on national honoraria. Medication costs for RSV outpatient episodes were estimated based on Belgian data from the ComNet study [3]. Full details are available in the KCE report[1].

*Equation 1: Cost per outpatient episode*

$$\text{Cost per outpatient episode} = \text{number of outpatient visits} \times (\%\ \text{paediatrician visits} \times \text{cost per paediatrician visit} + \%\ \text{GP visits} \times \text{cost per GP visit}) + \text{medication cost}$$

*Table S. 5 – Mean (95%CI) cost per outpatient episode in Belgium and in each region (based on equation 1)*

| **Cost per episode** | **Belgium** | **Flanders** | **Brussels** | **Wallonia** |
|---|---|---|---|---|
| **<1 year** | | | | |
| NIHDI | €77.43 (59.1-95.77) | €77.35 (59.5-95.21) | €86.75 (67.99-105.51) | €80.43 (61.33-99.53) |
| Patients | €28.58 (23.22-33.94) | €33.49 (28.37-38.61) | €27.61 (22.04-33.18) | €30.08 (24.33-35.82) |
| Total | €106.01 (82.32-129.71) | €95.54 (72.57-118.52) | €102.66 (78.32-126.99) | €110.51 (85.66-135.35) |
| **1 year** | | | | |
| NIHDI | €70.61 (53.55-87.66) | €73.01 (56.31-89.72) | €79.6 (62.24-96.97) | €72.77 (55.16-90.38) |
| Patients | €26.21 (21.49-30.93) | €29.55 (25.01-34.1) | €23.06 (18.18-27.93) | €27.29 (22.3-32.29) |
| Total | €96.82 (75.04-118.6) | €87.01 (65.75-108.26) | €94.62 (72.38-116.86) | €100.07 (77.46-122.67) |
| **2 years** | | | | |
| NIHDI | €67.76 (51.43-84.08) | €70.45 (54.4-86.49) | €76.49 (59.92-93.05) | €69.44 (52.69-86.2) |
| Patients | €24.78 (20.43-29.14) | €28.27 (24.05-32.49) | €21.5 (17.02-25.98) | €25.63 (21.06-30.2) |
| Total | €92.54 (71.86-113.22) | €83.15 (62.89-103.42) | €89.95 (68.9-110.99) | €95.07 (73.74-116.4) |
| **3 years** | | | | |
| NIHDI | €66.09 (50.19-81.99) | €68.95 (53.28-84.61) | €74.66 (58.56-90.75) | €67.52 (51.25-83.78) |
| Patients | €23.95 (19.81-28.09) | €27.52 (23.49-31.55) | €20.58 (16.34-24.83) | €24.67 (20.34-28.99) |
| Total | €90.04 (70-110.08) | €80.91 (61.21-100.6) | €87.2 (66.86-107.54) | €92.18 (71.59-112.77) |
| **4 years** | | | | |
| NIHDI | €65.14 (49.49-80.79) | €68.11 (52.66-83.57) | €73.61 (57.78-89.44) | €66.4 (50.42-82.38) |
| Patients | €23.48 (19.46-27.5) | €27.1 (23.18-31.02) | €20.06 (15.95-24.17) | €24.11 (19.92-28.29) |
| Total | €88.62 (68.94-108.29) | €79.66 (60.29-99.03) | €85.63 (65.69-105.57) | €90.51 (70.35-110.66) |

*95%CI: 95% confidence interval, NIHDI: National Institute for Health and Disability Insurance.*

##### 1.4.3 Non-medically attended (non-MA) episodes

The cost per non-medically attended episode was estimated using resource utilisation data reported by Belgian parents in the ComNet study [3], combined with unit costs of over-the-counter medications derived from an influenza-like illness study from the Belgian data of a multi-country influenza-like illness study [8].

##### 1.4.4 Cost associated with productivity losses

The percentage and mean number of workdays lost due to RSV outpatient and inpatient episodes were obtained from the ComNet study [3]. Per Belgian Pharmacoeconomics guideline [7], the average productivity cost per day was €376.8 based on the average labour costs including employee wages and/or salaries and employers' social security contributions in 2023 [9].

*Table S. 6 – Cost of parental productivity loss per RSV episode*

| Episode | Age | % lost workdays | Mean (SD) | Weighted mean cost per episode (SD) |
|---|---|---|---|---|
| Outpatient | <1y | 64% | 2.7 (3.3) days | € 636 (798) |
| | 1–4y | 80% | 4.3 (4.2) days | € 1,296 (1257) |
| Inpatient | <1y | 76% | 4.6 (4.4) days | € 1,318 (1261) |
| | 1–4y | 69% | 8.1 (10.7) days | € 2,107 (2790) |

*SD: standard deviation, Y: year.*

#### 1.5 Health-related quality-of-life

Using data from the RESCEU infant study, we recalculated QALY loss for infants using the Belgian EQ-5D-Y value set [10], and for caregivers using the EQ-5D-5L value set [11]. QALY results are shown in Table S. 7.

*Table S. 7 – Infant and their caregivers' quality-adjusted life-day (QALY) loss per RSV episode, stratified by health care resource utilisation*

| | | Mean (95%CI) | Median [IQR] | Sample size |
|---|---|---|---|---|
| Infant | Infant pooled | 0.00478 (0.0043 - 0.0053) | 0.00439 [0.00212 - 0.00689] | 180 |
| By healthcare resource utilisation | Outpatient | 0.00589 (0.0052 - 0.0067) | 0.00562 [0.00344 - 0.00744] | 81 |
| | Non-MA | 0.00337 (0.0028 - 0.004) | 0.00265 [0.00115 - 0.00491] | 90 |
| | Hospitalised | 0.00982 (0.0085 - 0.0114) | 0.00949 [0.00838 - 0.011] | 7 |
| Caregivers | Caregivers pooled | 0.00022 (-0.0001 - 0.0005) | 0.00008 [0 - 0.0008] | 164 |
| By healthcare resource utilization | Outpatient | 0.00051 (0.0002 - 0.0009) | 0.00033 [0 - 0.0009] | 75 |
| | Non-MA | -0.00028 (-0.0007 - 0.0001) | 0 [-0.00007 - 0.0003] | 81 |
| | Hospitalised | 0.00289 (0.0016 - 0.0045) | 0.00196 [0.00173 - 0.00375] | 6 |

*Non-MA: non-medical attendance, CI: confidence interval, IQR: interquartile range. In the cost-effectiveness analysis, we converted quality-adjusted life-day to quality-adjusted life-year.*

#### 1.6 Interventions' efficacy and durability

A systematic review of the literature on efficacy, effectiveness and safety of nirsevimab (NmAb) and maternal vaccine (MV) Abrysvo was conducted, with full details provided in the KCE report [1].

##### 1.6.1 Base case: efficacy values based on clinical trials.

In the base case, we used severity-specific efficacy estimates of RSV interventions as reported in randomised clinical trials (RCTs), assuming

- MA RSV-Lower Respiratory Tract Infection (LRTI) as a proxy for non-MA and outpatient RSV episode
- hospital admissions for RSV-LRTI as a proxy for hospital admissions with RSV
- severe MA RSV-LRTI as a proxy for RSV ICU admission and death (if reported)

For the RSV-preF MV, we used the final analysis by Simoes et al. (2025) [12], which confirmed the primary findings of Kampmann et al. (2023) [13]. Further details on incremental cases, sampling, and assumptions are provided in the KCE report [1]. Time-specific efficacy estimates with 95% credible intervals (CrI) were derived using a Bayesian approach, with results presented in Figure S. 5.

For nirsevimab, we assumed constant monthly efficacy over a 6-month period (Figure S. 6), based on 150-day efficacy reported in phase 2b and phase 3 RCTs [14-16], and supported by 180-day efficacy against hospitalisation observed in the phase 3b trial [17].

*Figure S. 5 – Estimated maternal vaccine efficacy values over 180 days (base case)*

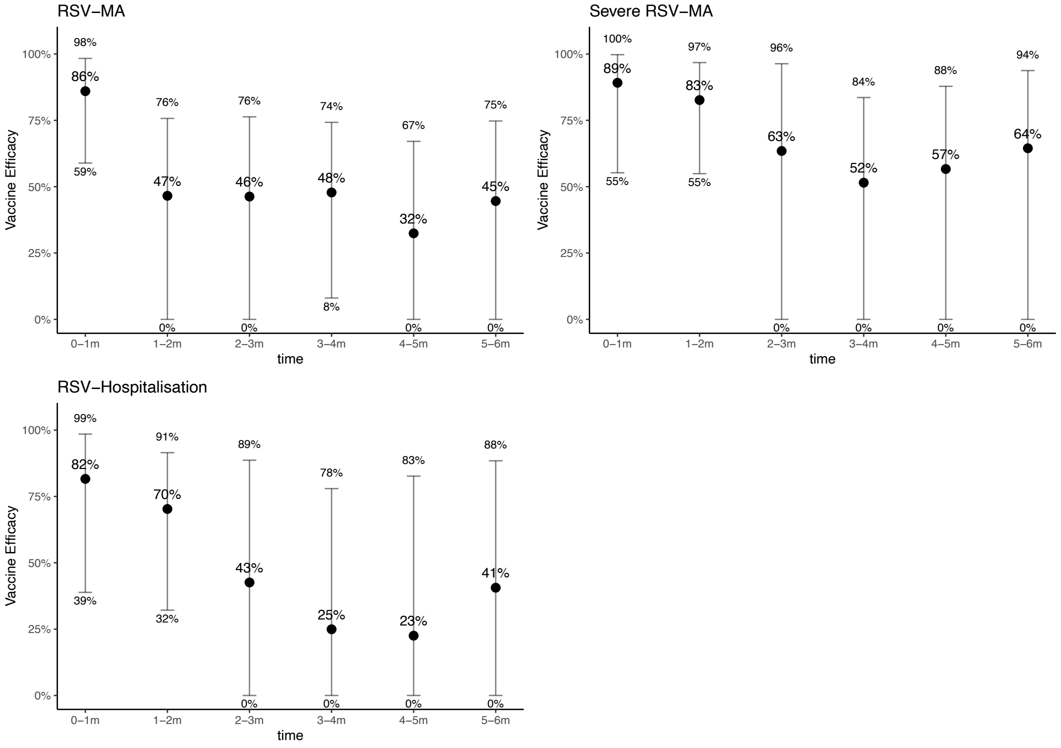


*m: month, MA: medically-attended cases.*

*Figure S. 6 – Reported nirsevimab efficacy value over 180 days (base case)*

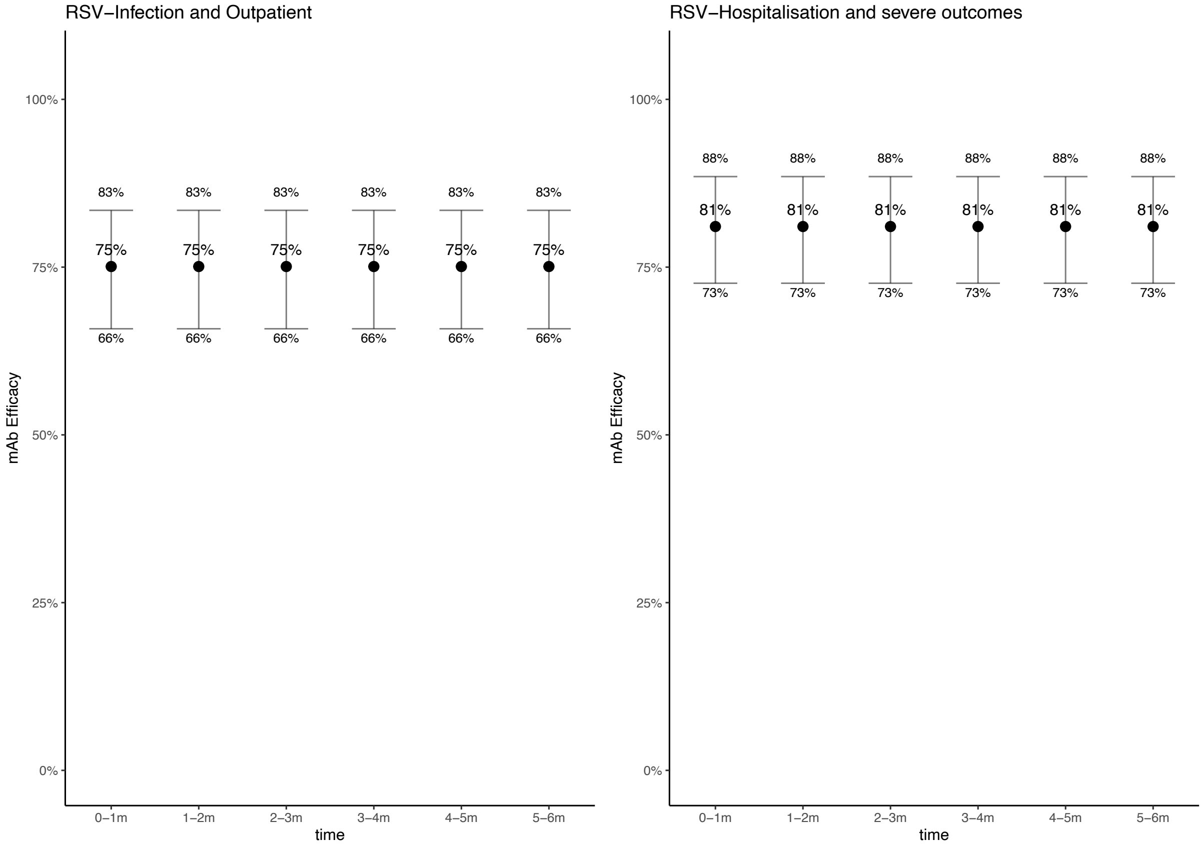


*m: month.*

##### *1.6.2 Scenario analysis: effectiveness based on real-world studies*

Scenario analyses also incorporated real-world effectiveness (RWE) studies of nirsevimab, including two sets of estimates: (1) pooled effectiveness from the KCE systematic review and meta-analysis [1], assuming constant protection over six months; and (2) a test-negative case-control study reporting effectiveness by time since immunisation [18].

*Table S. 8 – Scenario analyses based on two sets of nirsevimab effectiveness data*

| | **Nirsevimab (pooled real-world studies) with 6-month protection**<br>Mean and 95%CI | **Nirsevimab (US test-negative case-control study) with 5-month protection**<br>Mean[@] and 95%CI |
|---|---|---|
| Efficacy: non-MA episode | Assume same as an outpatient episode | 0-1 month: 76% (95%CI: 60-87%)<br>1-2 month: 70% (95%CI: 51-82%)<br>2-3 month: 61% (95%CI: 34-77%)<br>3-4 month: 47% (95%CI: 0-72%)[&]<br>4-5 month: 37% (95%CI: 0-70%)[&]<br>5-6 month: 0% |
| Efficacy vs. outpatient episode | 73% (95%CI: 67-78%)* | 0-1 month: 72% (95%CI: 48-86%)<br>1-2 month: 64% (95%CI: 38-80%)<br>2-3 month: 55% (95%CI: 20-76%)<br>3-4 month: 41% (95%CI: 0-70%)[&] |

| | | |
|---|---|---|
| | | 4-5 month: 32% (95%CI: 0-68%)[&]<br>5-6 month: 0% |
| Efficacy vs. hospitalisation | 86.0% (95%CI: 70-94%)[^] | 0-1 month: 89% (95%CI: 67-97%)<br>1-2 month: 83% (95%CI: 53-95%)<br>2-3 month: 74% (95%CI: 24-92%)<br>3-4 month: 59% (95%CI: 0-89%)[&]<br>4-5 month: 48% (95%CI: 0-88%)[&]<br>5-6 months: 0% |
| Efficacy vs. ICU admission and deaths | 87% (95%CI: 77-93%)* | 0-1 month: 94% (95%CI: 75-99%)<br>1-2 month: 88% (95%CI: 61-97%)<br>2-3 month: 78% (95%CI: 21-94%)<br>3-4 month: 58% (95%CI: 0-91%)[&]<br>4-5 month: 43% (95%CI: 0-89%)[&]<br>5-6 month: 0 |
| Duration of protection | 6 months protection | 5 months protection |

** Studies with follow-up superior to 100 days. ^ Studies with follow-up through 100 to 150 days. & All negative values were set to 0 in the analyses, and the distribution was truncated to align with the mean values. @ The original article from Xu et al. only reported median, the mean values were provided by the first author via personal communication. ICU: intensive care unit, MA: medically attended, CI: confidence interval, CrI: credible interval.*

#### 1.7 Long-term consequences

As described in a previous study (Li et al.[19] 2022), we evaluated the impact of recurrent wheezing (up to age 3) and the combined burden of wheezing and asthma (up to age 13) on cost-effectiveness outcomes for infants hospitalised with RSV in their first year of life. Annual probabilities of developing these conditions were based on six studies identified in two systematic reviews [20, 21] examining the link between early RSV infection and later respiratory morbidity (Table S. 9). We also intended to assess outcomes following RSV outpatient visits, but data were insufficient to estimate the probability of subsequent wheezing or asthma in non-hospitalised cases.

*Table S. 9 – Scenario analysis: probability of recurrent wheezing and asthma up to 13 years of age for infants who were hospitalised with RSV during their first year of life*

| **Scenario** | **Age (years)** | Annual probability of recurrent wheezing/asthma (uncertainty distribution) |
|---|---|---|
| Age 0–13 years | 0–1 | 0.31 ~ Beta (α =13, β =19) |
| | 1–2 | 0.27 ~ Beta (α =135, β =269) |
| | 2–3 | 0.17 ~ Beta (α =87, β =417) |
| | 3–4 | 0.16 ~ Beta (α =80, β =424) |
| | 4–5 | 0.10 ~ Beta (α =50, β =454) |
| | 5–13 | Same as age 4–5 |

Based on a Dutch study in preterm infants, the annual cost of recurrent wheezing included 5.5 primary care visits and one beta-agonist inhaler [20]. A Belgian study estimated medication costs for asthma-like symptoms in preschoolers at €42.72 (2014 value) [21]. Assuming typical prescribing patterns, we estimated that 10% of these costs were borne by patients.

*Table S. 10 – Undiscounted cost of recurrent wheezing and asthma per year used in scenario analysis*

| Cost per episode | Belgium | Flanders | Wallonia | Brussels |
|---|---|---|---|---|
| NIHDI | € 215.02 | € 212.41 | € 217.28 | € 219.13 |
| Patients | € 49.82 | € 48.57 | € 50.91 | € 51.80 |
| Healthcare payers | € 264.84 | € 260.98 | € 268.19 | € 270.93 |

*NIHDI: National Institute for Health and Disability Insurance.*

Due to the lack of Belgian HRQoL data for children with asthma, we used pooled EQ-5D-Y data from studies in Sweden [22], Spain [23], and the Netherlands [24]. Applying the Belgian value set, we estimated a health utility of 0.9217, implying an annual QALY loss of 0.0183 compared to the population norm of 0.94 [25].

Overall, the total undiscounted treatment costs and QALY losses associated with recurrent wheezing among children hospitalised with RSV during their first year of life were calculated as:

$$\text{Total Cost} = \text{Nr of hospitalisations}_{\text{(age 0-11 months)}} \times P_{\text{(year)}} \times \text{Annual treatment cost}$$

$$\text{QALY loss} = \text{Nr of hospitalisations}_{\text{(age 0-11 months)}} \times P_{\text{(year)}} \times \text{QALY loss per hospitalised case}$$

Where $P_{year}$ represents the probability of recurrent wheezing and asthma at a given age (Table S. 11).

Full methodology are detailed in the KCE report, titled "Cost-effectiveness of new preventive options against RSV infections in Belgian infants" [1].

### 2 Supplement results

#### 2.1 RSV-related disease and economic burden in Belgian children under 5 years

In Belgium, RSV led to a substantial disease burden in children under 5 years of age, as illustrated in Table S. 11.

*Table S. 11 – Estimated RSV-related undiscounted burden without new RSV intervention in a birth cohort followed over 5 years (Mean [95%CrI])*

| Outcome | 0–2 months | 3–5 months | 6–11 months | 0–11 months | 12–59 months | 0–59 months |
|---|---|---|---|---|---|---|
| Non-MA episodes | 2,419 [1,282 ; 4,472] | 3,598 [1,709 ; 6,947] | 4,091 [1,923 ; 8,500] | 10,108 [4,944 ; 19,956] | 30,373 [11,712 ; 68,439] | 40,481 [16,594 ; 88,731] |
| Outpatient episodes | 5,593 [1,951 ; 12,095] | 6,524 [2,457 ; 12,958] | 6,015 [2,139 ; 13,362] | 18,132 [6,544 ; 38,415] | 48,117 [17,110 ; 106,898] | 66,249 [23,654 ; 145,314] |
| Hospitalisations (non-ICU) | 3,169 | 1,641 | 1,519 | 6,329 | 2,309 | 8,638 |
| ICU admissions | 245 | 72 | 67 | 384 | 43 | 428 |
| **Total cases** | 11,427 [6,805 ; 19,850] | 11,835 [6,018 ; 22,074] | 11,692 [5,764 ; 23,583] | 34,954 [18,587 ; 66,019] | 80,842 [31,456 ; 179,436] | 115,796 [50,026 ; 245,605] |
| Deaths | 2.0 | 0.9 | 0.8 | 3.7 | 1.3 | 5.0 |
| Life years lost | 166 | 72 | 66 | 304 | 108 | 411 |
| **QALY** | | | | | | |
| QALY loss due to non-MA episodes | 8.1 [4.2 ; 15] | 12 [5.6 ; 24] | 14 [6.2 ; 29] | 34 [16 ; 68] | 102 [38 ; 227] | 136 [54 ; 294] |
| QALY loss due to outpatient episodes | 33 [11 ; 72] | 38 [15 ; 78] | 35 [13 ; 78] | 107 [39 ; 227] | 283 [102 ; 623] | 390 [140 ; 851] |
| QALY loss due to hospitalisation | 31 [27 ; 36] | 16 [14 ; 19] | 15 [13 ; 17] | 62 [53 ; 72] | 23 [19 ; 26] | 85 [73 ; 98] |
| QALY loss due to ICU admission | 2.4 [2.1 ; 2.8] | 0.7 [0.6 ; 0.8] | 0.7 [0.6 ; 0.8] | 3.8 [3.2 ; 4.4] | 0.4 [0.4 ; 0.5] | 4.2 [3.6 ; 4.8] |
| QALY loss due to deaths | 142 | 61 | 57 | 261 | 92 | 353 |
| **Total QALY loss** | 217 [192 ; 261] | 129 [99 ; 178] | 121 [91 ; 177] | 467 [380 ; 617] | 500 [257 ; 950] | 968 [640 ; 1,561] |
| **Cost (€'000)** | | | | | | |
| Cost due to non-MA episodes | 11 [5.7 ; 20] | 16 [7.6 ; 31] | 18 [8.5 ; 38] | 45 [22 ; 88] | 135 [52 ; 303] | 179 [74 ; 393] |
| Cost due to outpatient episodes | 593 [208 ; 1,347] | 692 [267 ; 1,455] | 629 [226 ; 1,455] | 1,915 [696 ; 4,263] | 4,425 [1,589 ; 9,887] | 6,340 [2,240 ; 14,049] |
| Cost due to hospitalisations (non-ICU) | 11,451 [11,374 ; 11,527] | 5,929 [5,889 ; 5,968] | 5,418 [5,386 ; 5,451] | 22,797 [22,646 ; 22,945] | 7,438 [7,355 ; 7,525] | 30,235 [30,070 ; 30,415] |
| Cost due to ICU admission | 3,502 [3,343 ; 3,651] | 1,035 [988 ; 1,079] | 986 [947 ; 1,027] | 5,524 [5,285 ; 5,757] | 767 [675 ; 865] | 6,291 [6,039 ; 6,548] |
| **Total treatment cost** | 15,557 [15,121 ; 16,299] | 7,672 [7,238 ; 8,441] | 7,051 [6,633 ; 7,902] | 30,280 [29,031 ; 32,614] | 12,764 [9,858 ; 18,304] | 43,045 [38,864 ; 50,994] |

*Grey area: the total columns (0–11 months and 0–59 months), the totals may differ by up to one unit due to rounding. QALY: quality-adjusted life year, non-MA: non-medically attended, ICU: intensive care unit, CrI: credible interval.*

#### 2.2 Impact of interventions on RSV disease and economic burden

*Table S. 12 – Disease and economic burden averted in children <5 years compared to 'no intervention' from the HCP perspective (Mean [95%CrI])*

| | MV | MV: Sep-Mar | NmAb | NmAb: Oct-Mar | NmAb: Oct-Mar + catch-up |
|---|---|---|---|---|---|
| *Coverage* | *40%* | *40%* | *90%* | *90%* | *90%* |
| **Undiscounted cases averted** | | | | | |
| Non-MA episodes | 1,069 [321 ; 2,267] | 621 [194 ; 1,297] | 4,039 [2,009 ; 7,713] | 1,452 [770 ; 2,692] | 5,222 [2,583 ; 10,125] |
| Outpatient episodes | 2,439 | 1,620 | 8,127 | 3,528 | 9,845 |

| | | | | | |
|---|---|---|---|---|---|
| | [678 ; 5,578] | [450 ; 3,688] | [2,950 ; 16,666] | [1,220 ; 7,491] | [3,554 ; 20,302] |
| Hospitalisations (non-ICU) | 971 [412 ; 1,567] | 796 [359 ; 1,196] | 3,494 [3,085 ; 3,812] | 1,966 [1,735 ; 2,144] | 4,062 [3,586 ; 4,431] |
| ICU admissions | 93 [51 ; 119] | 80 [45 ; 99] | 230 [203 ; 251] | 160 [141 ; 175] | 252 [223 ; 275] |
| Total cases averted | 4,572 [1,941 ; 8,904] | 3,116 [1,423 ; 5,776] | 15,890 [8,883 ; 28,088] | 7,106 [4,191 ; 12,473] | 19,381 [10,703 ; 34,815] |
| Deaths | 0.8 [0.4 ; 1.1] | 0.7 [0.4 ; 0.8] | 2.1 [1.9 ; 2.3] | 1.3 [1.1 ; 1.4] | 2.4 [2.1 ; 2.6] |
| Life-years lost | 66 [36 ; 88] | 53 [30 ; 68] | 173 [152 ; 188] | 105 [93 ; 115] | 197 [174 ; 214] |
| **Discounted QALYs gained (rate 1.5%)** | | | | | |
| QALYs gained due to non-MA episodes | 3.6 [1.0 ; 7.7] | 2.1 [0.6 ; 4.5] | 13.5 [6.4 ; 26.4] | 4.9 [2.4 ; 9.4] | 17.5 [8.3 ; 34.4] |
| QALYs gained due to outpatient episodes | 14.4 [3.9 ; 33] | 9.6 [2.6 ; 22] | 47.9 [17.3 ; 101] | 20.8 [7.2 ; 45] | 58.0 [20.8 ; 124] |
| QALYs gained due to hospitalisations | 9.6 [4.0 ; 16] | 7.8 [3.6 ; 12] | 34.4 [28.2 ; 40] | 19.3 [15.9 ; 23] | 39.9 [32.8 ; 47] |
| QALYs gained due to ICU admission | 0.9 [0.5 ; 1.2] | 0.8 [0.4 ; 1.0] | 2.3 [1.9 ; 2.7] | 1.6 [1.3 ; 1.9] | 2.5 [2.0 ; 2.9] |
| QALYs gained due to deaths | 33 [18 ; 44] | 27 [15 ; 34] | 86 [76 ; 94] | 53 [46 ; 57] | 98 [87 ; 107] |
| **Total discounted QALYs gained** | 61 [40 ; 88] | 47 [31 ; 65] | 184 [145 ; 248] | 99 [81 ; 127] | 216 [168 ; 296] |
| **Discounted treatment costs saved (€ '000) (rate 3%)** | | | | | |
| Direct cost saved due to non-MA episodes | 4.7 [1.4 ; 10.0] | 2.8 [0.9 ; 5.8] | 17.9 [8.9 ; 34.2] | 6.4 [3.4 ; 11.9] | 23.1 [11.4 ; 44.9] |
| Direct cost saved due to outpatient episodes | 259 [71 ; 620] | 172 [49 ; 409] | 862 [317 ; 1,893] | 374 [132 ; 843] | 1,045 [385 ; 2,315] |
| Direct cost saved due to hospitalisations | 3,508 [1,488 ; 5,653] | 2,875 [1,298 ; 4,329] | 12,625 [11,138 ; 13,759] | 7,102 [6,265 ; 7,739] | 14,675 [12,946 ; 15,992] |
| Direct cost saved due to ICU admissions | 1,330 [730 ; 1,701] | 1,139 [652 ; 1,421] | 3,296 [2,912 ; 3,641] | 2,289 [2,022 ; 2,528] | 3,609 [3,189 ; 3,986] |
| **Total treatment cost averted** | 5,102 [2,986 ; 7,271] | 4,189 [2,547 ; 5,683] | 16,802 [14,792 ; 18,466] | 9,772 [8,638 ; 10,689] | 19,351 [17,013 ; 21,315] |
| **Intervention and incremental costs (€ '000)** | | | | | |
| **Intervention costs (list price*)** | 8,086 | 4,717 | 76,057 | 38,028 | 76,057 |
| **Incremental costs** | 2,984 [815; 5,5100] | 528 [-996 ; 2,572] | 59,255 [57,591 ; 61,264] | 28,257 [27,339 ; 29,390] | 56,706 [54,742 ; 59,044] |

*Negative incremental costs indicate savings. * The cost of the intervention includes the cost per dose of the product (valued at list price), excluding delivery costs. CrI: credible interval, QALY: quality-adjusted life year, non-MA: non-medically attended, ICU: intensive care unit, MV: year-round maternal vaccine, MV: Sep-Mar: seasonal maternal vaccine from September to March, NmAb: year-round nirsevimab, NmAb: Oct-Mar: seasonal NmAb strategy from October to March, NmAb: Oct-Mar + catch-up: seasonal NmAb combined with a catch-up strategy.*

#### 2.3 Cost-effectiveness of RSV immunisation strategies

##### *2.3.1 Cost-effectiveness of each strategy versus standard of care*

Table S. 13 presents the incremental cost-effectiveness ratios (ICERs) for each RSV immunisation strategy compared to no intervention. Using current list prices, all 3 NmAb strategies exceeded the €250,000 per QALY willingness-to-pay threshold. Under the cost-parity scenario, however, the seasonal NmAb strategy was broadly cost-saving, and the seasonal plus catch-up NmAb strategy had an ICER below €1,000 per QALY gained compared to no intervention.

*Table S. 13 – Expected incremental cost-effectiveness ratios of each strategy compared to 'no intervention' from the HCP perspective (Mean [95%CrI])*

| | **MV** | **MV: Sep-Mar** | **NmAb** | **NmAb: Oct-Mar** | **NmAb: Oct-Mar + catch-up** |
|---|---|---|---|---|---|
| Total discounted QALYs gained | 61.40 [39.77 ; 87.51] | 46.86 [30.84 ; 64.61] | 184.24 [144.82 ; 247.86] | 99.08 [80.79 ; 126.91] | 216.08 [167.96 ; 296.14] |

| | | | | | |
|---|---|---|---|---|---|
| Total discounted treatment cost averted (€'000) | 5,102 [2,986 ; 7,271] | 4,189 [2,548 ; 5,683] | 16,802 [14,792 ; 18,466] | 9,772 [8,638 ; 10,689] | 19,351 [170,013 ; 21,315] |
| **Base case: using list prices** | | | | | |
| Intervention costs at list price (€'000) | 8,086 | 4,717 | 76,057 | 38,028 | 76,057 |
| Incremental costs (€'000) | 2,984 [815; 5,100] | 528 [-966; 2,170] | 59,255 [57,591 ; 61,264] | 28,257 [27,339; 29,390] | 56,706 [54,742; 59,044] |
| ICER per QALY gained | 48,607 | 11,276 | 321,614 | 285,190 | 262,422 |
| **Cost parity scenario: €200 per dose for both interventions including delivery costs** | | | | | |
| Intervention costs (€'000) | 8,694 | 5,072 | 19,562 | 9,781 | 19,562 |
| Incremental costs (€'000) | 3,593 [1,423 ; 5,708] | 883 [-611 ; 2,524] | 2,760 [1096 ; 4,770] | 9 [-908 ; 1,143] | 211 [-1,752 ; 2,549] |
| ICER / QALY gained versus no intervention | 58,513 | 18,846 | 14,982 | 96 | 978 |

*Negative incremental costs indicate savings, QALY: quality adjusted life-year, CrI: credible interval, ICER: incremental cost-effectiveness ratio, MV: year-round maternal vaccine, MV: Sep-Mar: seasonal maternal vaccine from September to March, NmAb: year-round nirsevimab, NmAb: Oct-Mar: seasonal NmAb strategy from October to March, NmAb: Oct-Mar + catch-up: seasonal NmAb combined with a catch-up strategy.*

##### 2.3.2 Incremental cost-effectiveness plane: full incremental analysis

A full incremental analysis compared all 5 RSV strategies to no intervention and each other. The ICERs, excluding dominated strategies, are reported in Table S. 14. In the base case with list prices, the seasonal MV strategy had an ICER of €11,276 per QALY gained (vs no intervention), while the NmAb seasonal plus catch-up strategy had the highest ICER (€347,290 vs year-round MV). Under cost parity, only the seasonal and seasonal plus catch-up NmAb strategies remained on the frontier, with ICERs of €96 (vs no intervention) and €1,725 per QALY gained (vs seasonal NmAb), respectively. These results align with the findings in Table S. 13 and the cost-effectiveness plane shown in Figure 2 (main text).

*Table S. 14 – Expected incremental cost-effectiveness ratios of the full incremental analysis (HCP perspective, versus the next best alternative)*

| **Strategies** | **Strategies compared to** | **Incremental cost ('000)** | **Incremental QALY** | **ICER per QALY gained** |
|---|---|---|---|---|
| **Base case: list prices** | | | | |
| MV: Sep-Mar | 'no intervention' | 528 [-966 ; 2,170] | 46.86 [30.84 ; 64.61] | € 11,276 |
| MV | MV: Sep-Mar | 2,456 [1,758 ; 2,965] | 14.54 [8.11 ; 23.29] | € 168,938 |
| NmAb | NA | NA | NA | Dominated |
| NmAb: Oct-Mar | NA | NA | NA | Extendedly dominated |
| NmAb: Oct-Mar + catch-up | MV | 53,721 [50,694 ; 56,695] | 154.69 [111.38 ; 222.01] | € 347,290 |
| **Scenario: cost-parity** | | | | |
| MV: Sep-Mar | NA | NA | NA | Extendedly dominated |
| MV | NA | NA | NA | Extendedly dominated |
| NmAb | NA | NA | NA | Extendedly dominated |
| NmAb: Oct-Mar | 'no intervention' | 9 [-908; 1,143] | 99.08 [80.79 ; 126.91] | €96 |
| NmAb: Oct-Mar + catch-up | NmAb: Oct-Mar | 202 [-1,752; 2,549] | 117.00 [86.15 ; 169.22] | € 1,725 |

*HCP: health care payers, QALY: quality adjusted life-year, ICER: incremental cost-effectiveness ratio, MV: year-round maternal vaccine, MV: Sep-Mar: seasonal maternal vaccine from September to March, NmAb: year-round nirsevimab, NmAb: Oct-Mar: seasonal NmAb strategy from October to March, NmAb: Oct-Mar + catch-up: seasonal NmAb combined with a catch-up strategy.*

*Figure S. 7 – Cost-effectiveness analysis from the HCP perspective over a range of WTP threshold*

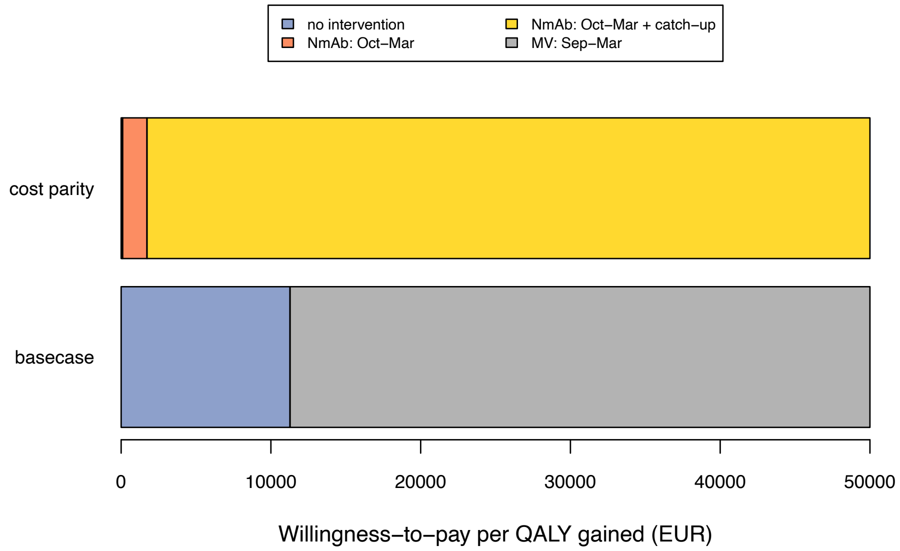


*values*

*WTP: willingness-to-pay, HCP: health care payers', EUR: euro, QALY: quality-adjusted life-year. MV: Sep-Mar: seasonal maternal vaccine from September to March, NmAb: Oct-Mar: seasonal nirsevimab strategy from October to March, NmAb: Oct-Mar + catch-up: seasonal NmAb combined with a catch-up strategy.*

##### *2.3.3 A full incremental analysis compared all five RSV strategies to no intervention and each other; ICERs excluding dominated strategies are reported in*

In addition to the cost-effectiveness plane, the CEACs (Figure S. 8, left) display the probability of each strategy being cost-effective across WTP values. ENLCs (Figure S. 8, right) identify the cost-effective strategy over a range of WTP values per QALY, showing the strategy with the lowest expected net loss.

*Figure S. 8 – CEACs (left plots) and ENLCs (right plots) comparing 5 RSV immunisation strategies in children from the HCP perspective*

**Base case: list price costs (MV: €186.01 and NmAb: €777.58 assumed to include delivery costs)**

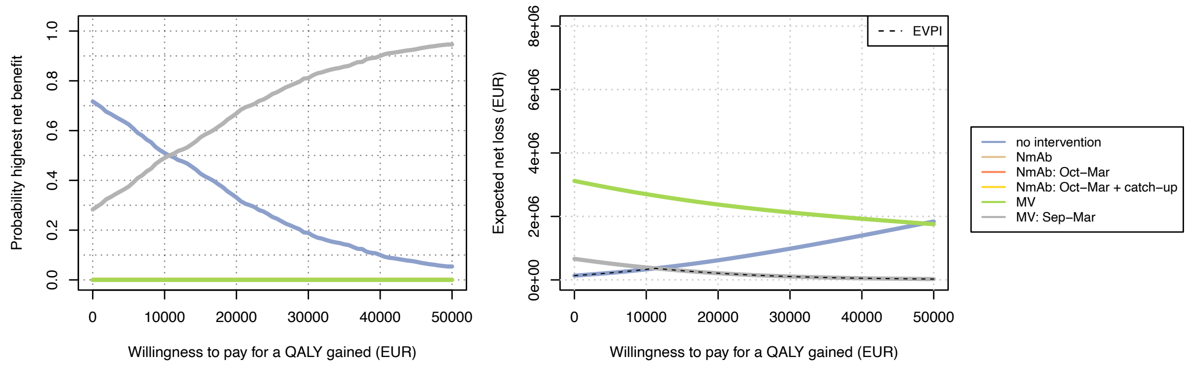


**Scenario: cost parity (€200 per dose, assumed to include delivery cost)**

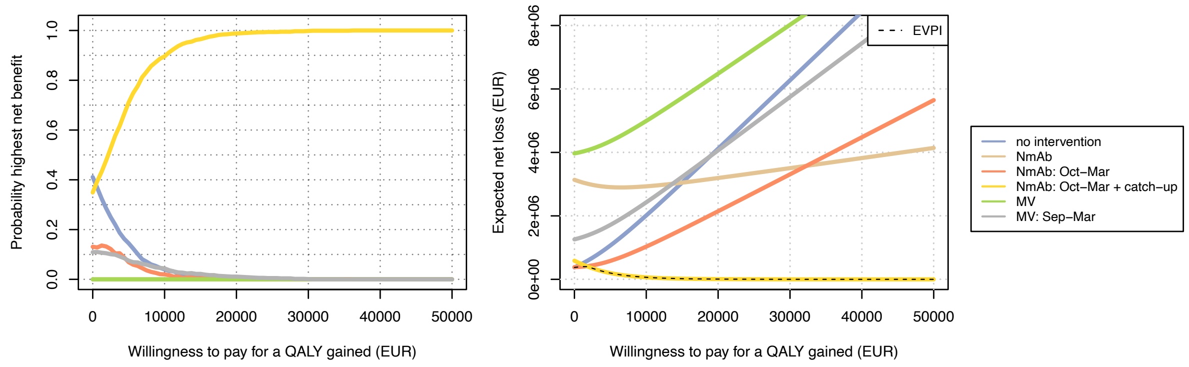


*CEAC: cost-effectiveness acceptability curve, ENLC: expected net loss curve, MV: year-round maternal vaccine, MV: Sept-Mar: seasonal maternal vaccine from September to March, NmAb: year-round nirsevimab, NmAb: Oct-Mar: seasonal NmAb strategy from October to March, NmAb: Oct-Mar + catch-up: seasonal NmAb combined with a catch-up strategy.*

##### *2.3.4 Bivariant threshold analysis of interventions' cost*

The seasonal NmAb strategy was the preferred strategy only over a narrow NmAb cost per dose range and given a WTP value of €0, €20,000 and €50,000 per QALY gained without and with the 'combined' strategy (Figure S. 9 and Figure S. 10).

*Figure S. 9 – Intervention cost threshold analysis from the HCP perspective (all-inclusive cost per dose), comparing 5 strategies to no intervention and to each other at willingness to pay of €0, €20,000 and €50,000 per QALY gained*

**Willingness to pay: €0 per QALY gained**

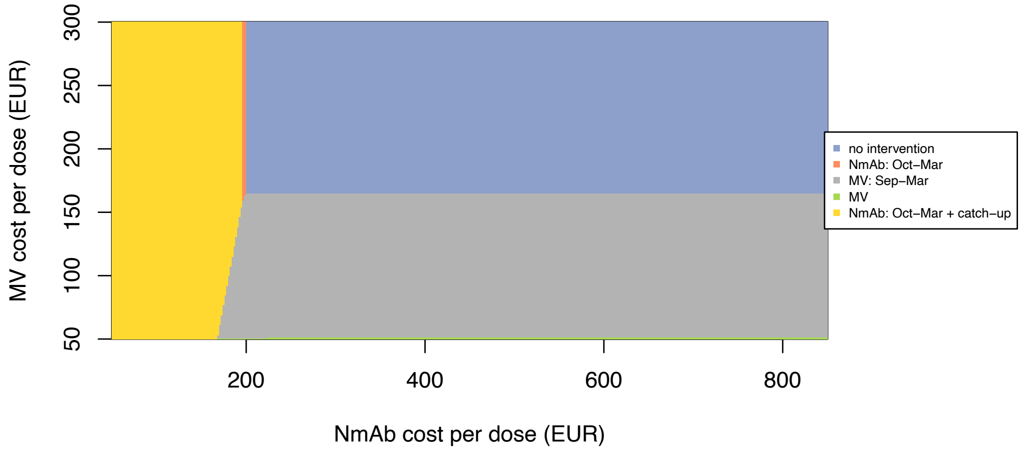


**Willingness to pay: €20,000 per QALY gained**

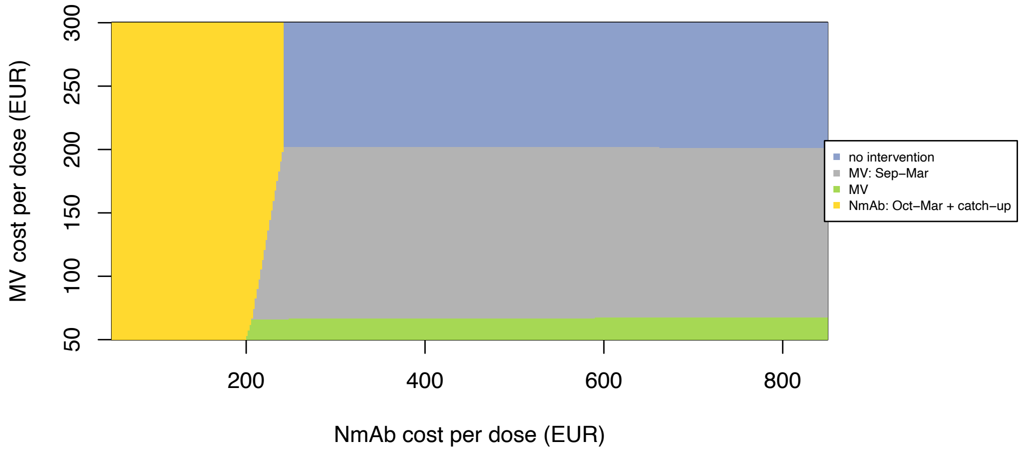


**Willingness to pay: €50,000 per QALY gained**

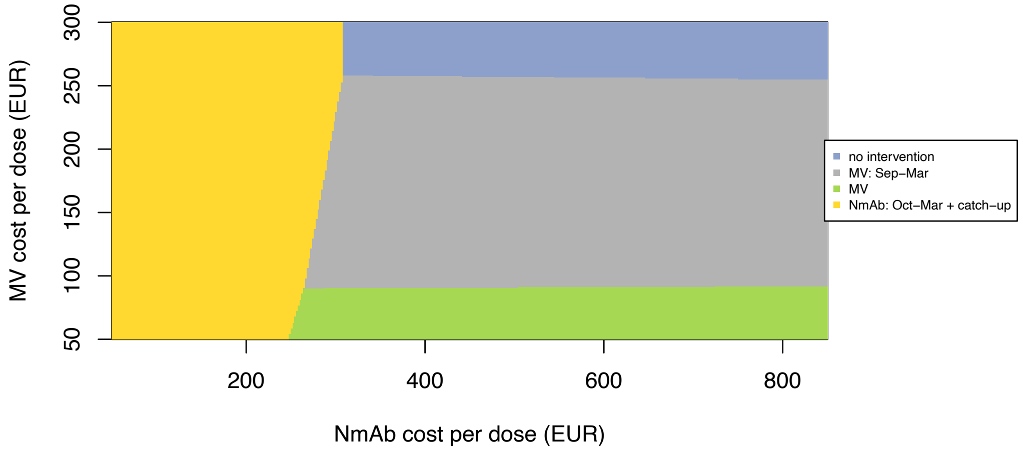


*Y-axis: maternal vaccine costs ranged from €50-€30, X-axis: NmAb costs from €50–€850. Each colour indicates a preferred RSV strategy, with the greatest uncertainty at the boundaries where strategies change. EUR: euro, HCP: health care payers', QALY: quality-adjusted life-year, MV: year-round maternal vaccine, MV: Sept-Mar: seasonal maternal vaccine from September to March, NmAb: Oct-Mar: seasonal NmAb strategy from October to March, NmAb: Oct-Mar + catch-up: seasonal NmAb combined with a catch-up strategy.*

*Figure S. 10 – Intervention cost threshold analysis from the HCP perspective (all-inclusive cost per dose), with a 'combined' strategy at willingness to pay of €0, €20,000 and €50,000 per QALY gained*

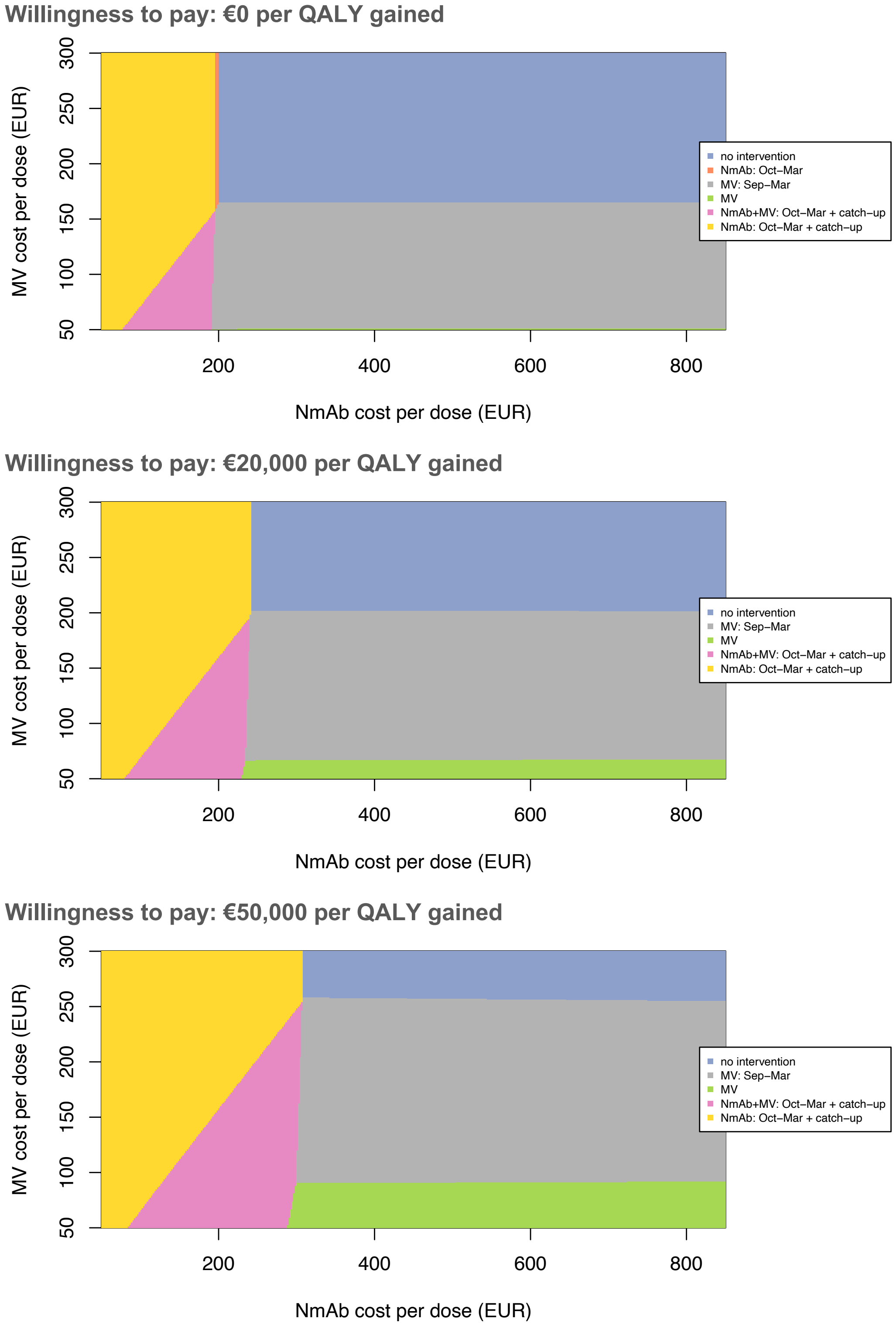


*Y-axis: maternal vaccine costs ranged from €50-€30, X-axis: NmAb costs from €50–€850. Each colour indicates a preferred RSV strategy, with the greatest uncertainty at the boundaries where strategies change. EUR: euro, HCP: health care payers', QALY: quality-adjusted life-year, MV: year-round maternal vaccine, MV: Sept-Mar: seasonal maternal vaccine from September to March, NmAb: Oct-Mar: seasonal NmAb strategy from October to March, NmAb: Oct-Mar + catch-up: seasonal NmAb combined with a catch-up strategy, NmAb + MV: Oct-Mar + catch-up: combined strategy: seasonal maternal vaccine from September to March, seasonal NmAb strategy from October to March for infants not immunised, and a catch-up NmAb strategy.*

##### *2.3.5 Expected value of partial perfect information*

The EVPPI was estimated for each uncertain input across varying WTP thresholds to identify key drivers of decision uncertainty. As shown in Figure S. 11, higher EVPPI values indicate greater influence of the corresponding parameter. Using both list prices and cost parity, EVPPI peaked at WTP values of around €11,000 and €1,000 per QALY, respectively, indicating greatest decision uncertainty at thresholds where preferred strategies change.

Overall, research that would reduce the uncertainty in our analysis the most, would be on estimating the interventions' efficacies against severe RSV outcomes and RSV outpatient burden.

*Figure S. 11 – Expected value of partial perfect information*

**Base case: list prices (MV: €186.01 and NmAb: €777.58 assumed to include delivery costs)**

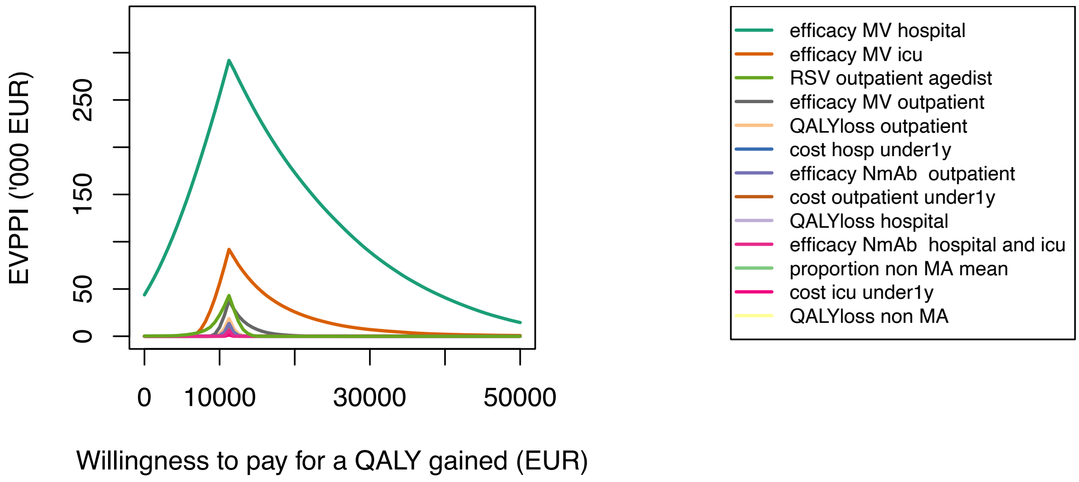


**Scenario: cost parity (€200 per dose, assumed to include delivery cost)**

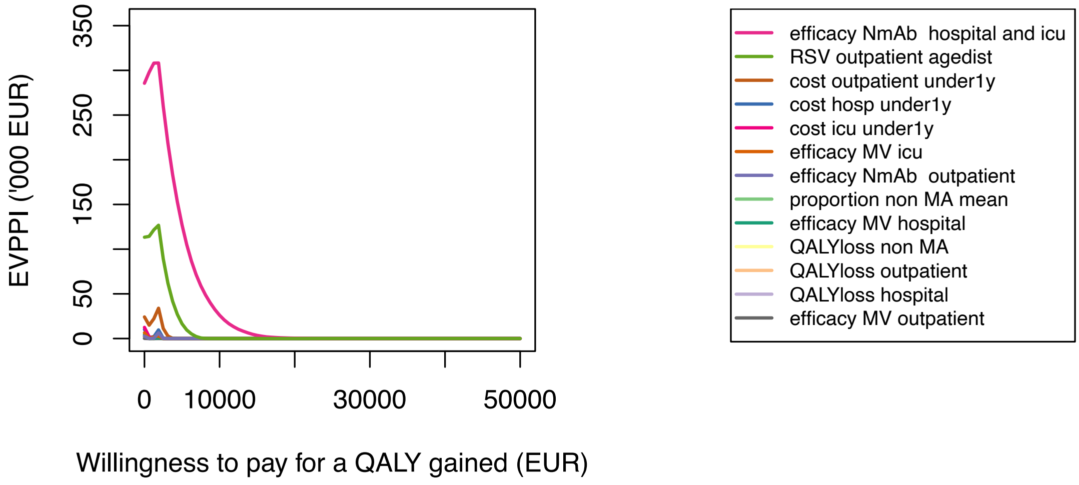


*EUR: euro, QALY: quality-adjusted life-year, EVPPI: expected value of partial perfect information, agedist: combined uncertainty range for age-specific RSV outpatient incidence rate, including age group: 0-2 months, 3-5 months and 6-11 months, MA: medically attendance, MV: maternal vaccine, icu: intensive care unit, NmAb: nirsevimab.*

##### 2.3.6 Scenario analyses

A list of scenario analyses was summarised in Table S. 15. The results of scenario analyses are presented in Figure S. 12 and Figure 6 in the main text. Additional bivariant price threshold analyses of each scenario are presented in KCE report [1].

Figure S. 12 compares the cost-effectiveness of RSV strategies from the societal and HCP perspectives. Using list prices, the seasonal MV strategy became preferred at a lower WTP under the societal perspective due to productivity cost savings. Under cost parity (€200/dose), the seasonal plus catch-up NmAb strategy was cost-saving from the societal perspective.

*Table S. 15 – List of scenario analyses*

| **Label in the graphs** | **Full name** | **Comparison of Base Case and Scenario Analyses (Differing from Base Case Assumptions)** |
|---|---|---|
| ***Base case*** | Base case analysis | • Perspective: HCP<br>• Price: list price (no delivery cost)<br>• RSV-coded hospitalisations: both primary and secondary diagnosis codes<br>• TCT data: average 4 seasons: 2016/2017 – 2019/2020<br>• Age distribution in 1-11m age group: season 2018/2019<br>• NmAb's efficacy values: pooled phase 3 RCT data, constant protection over 6 months<br>• Both interventions' efficacies against mortality: assume same as efficacies against hospitalisation (MV) or ICU (NmAb)<br>• Both interventions' efficacies against non-MA: assume same as efficacies against outpatient<br>• Inclusion of recurrent wheezing: no<br>• QALY losses: children only |
| ***Cost parity*** | SA: cost parity | • Cost: €200 per dose for either NmAb or MV (including all delivery costs) |
| ***Societal perspective*** | SA: Societal perspective | • Perspective: societal |
| ***Include 'combined' strategy*** | SA: Include 'combined' strategy' | • Added an additional strategy: *seasonal maternal vaccine from September to March, seasonal NmAb strategy from October to March for infants not immunised, and a catch-up NmAb strategy* |
| ***Hospitalisation data related scenarios*** | | |
| ***Hosp (a): Average 10 seasons*** | SA: using the average of 10 seasons non-ICU and ICU admissions data | • TCT data: average 10 seasons: 2008/2009 - 2013/2014 and 2016/2017 – 2019/2020 (see Figure S. 1) |
| ***Hosp (b): ICD primary code only*** | SA: using only the primary diagnosis code | • RSV-coded hospitalisations: using only the primary diagnosis code to select hospitalisations |
| ***Hosp (c):S23to24 age distribution*** | SA: using season 2023/2024 age distribution data | • Age distribution in 1-11m age group: season 2023/2024 |
| ***Hosp (d): S23to24 age primary code only*** | SA: using season 2023/2024 age distribution data and primary diagnosis code | • RSV-coded hospitalisations: using only the primary diagnosis code to select hospitalisations<br>• Age distribution in 1-11m age group: season 2023/2024 |
| ***Outpatient data related scenarios*** | | |
| ***OP incidence: NLD*** | SA: using the outpatient incidence rate from the Netherlands | • RSV-related primary care incidence: based on Dutch data |
| ***OP incidence: pooled (5 countries)*** | SA: using the pooled outpatient incidence rates | • RSV-related primary care incidence: based on pooled estimates from 5 countries |

| | | |
|---|---|---|
| ***Interventions' efficacy/effectiveness related scenarios*** | | |
| ***Efficacy NmAb RWE (6m constant)*** | SA: effectiveness data of nirsevimab using RWE studies | • NmAb's effectiveness values: pooled effectiveness data, constant protection over 6 months, 0% from month 7 onwards |
| ***Efficacy NmAb wane over 5m*** | SA: effectiveness data of nirsevimab using a test-negative case-control study | • NmAb's effectiveness values: wane over 5 months, 0% from month 6 onwards |
| ***No efficacy against death*** | SA: no protection against RSV mortality | • Both interventions' efficacies against mortality: assume no protection against RSV mortality |
| ***No efficacy against nonMA*** | SA: no protection against RSV non-MA episode | • Both interventions' efficacies against non-MA: assume no protection against RSV non-MA episode |
| ***Asthma wheezing up to 3y*** | SA: inclusion of recurrent wheezing and asthma up to 3 years | • Inclusion of recurrent wheezing and asthma up to 3 years of age in children who had an RSV hospitalisation before age 1 year |
| ***Asthma wheezing up to 13y*** | SA: inclusion of recurrent wheezing and asthma up to 13 years | • Inclusion of recurrent wheezing and asthma up to 13 years of age in children who had an RSV hospitalisation before age 1 year |
| ***Health outcome related scenario*** | | |
| ***QALY losses of caregivers*** | SA: inclusion of parental QALY losses per RSV episode | • Inclusion of parental QALY losses per RSV episode in addition to the children's QALY losses |

*SA: scenario analysis, HCP: health care payers', QALY: quality adjusted life-year, y: year, m: month, ICD: international classification of diseases, NmAb: nirsevimab, non-MA: non-medically attended, OP: outpatient, RWE: real-world evidence, S: season, NLD: the Netherlands.*

*Figure S. 12 – Scenario analyses related to the societal perspective*

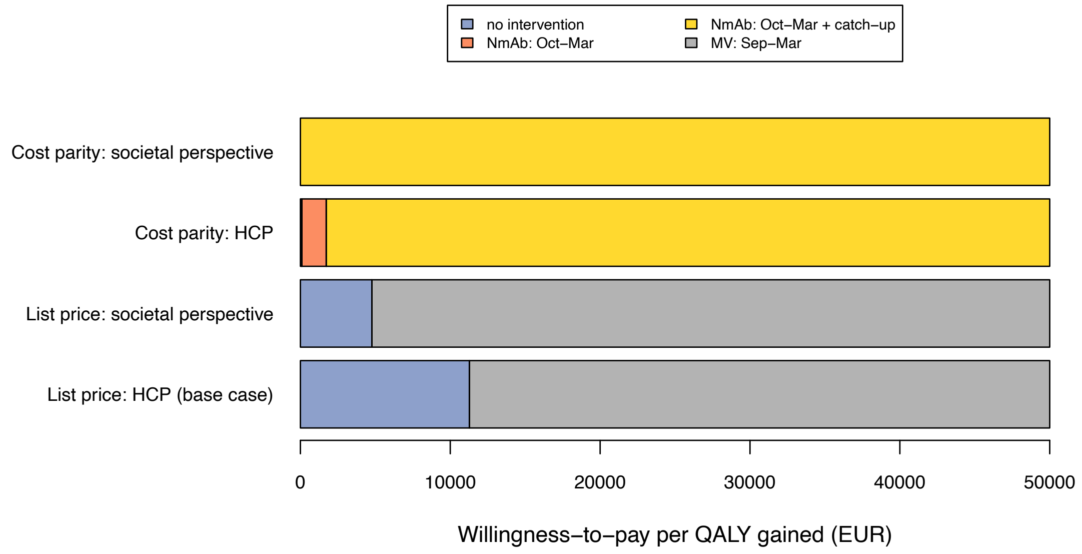


*EUR: euro, HCP: health care payers', QALY: quality-adjusted life-year, MV: Sept-Mar: seasonal maternal vaccine from September to March, NmAb: Oct-Mar: seasonal NmAb strategy from October to March, NmAb: Oct-Mar + catch-up: seasonal NmAb combined with a catch-up strategy.*

Additional scenario-specific bivariate threshold results are detailed in KCE report, titled "Cost-effectiveness of new preventive options against RSV infections in Belgian infants"[1].

#### 2.4 Budget impact analysis

BIA results for MV and NmAb strategies are shown in Table S. 16 and Table S. 17, with national-level data; demographic breakdowns by federated entity (Flanders, Brussels, and Wallonia) are available in the KCE report [1].

Under base case assumptions, year-round and seasonal MV strategies at 40% coverage avoided €5.1 million and €4.2 million in treatment costs, respectively (Table S. 16). Assuming €50–€250 per dose, immunisation costs ranged from €2.1–€10.9 million (year-round) and €1.3–€6.3 million (seasonal). At €50 per dose, RoI was highest: €2.40 per €1 invested for year-round MV and €3.30 for seasonal MV. At €150 per dose, year-round MV nearly broke even (RoI 0.78), while seasonal MV still yielded a 10% return (RoI 1.10). Seasonal MV consistently offered a more favourable RoI across all cost assumptions.

At 90% coverage, the seasonal NmAb and seasonal plus catch-up NmAb strategies were estimated to reduce treatment costs by €9.8 million and €19.4 million, respectively (Table S. 17). Assuming all-inclusive costs of €50–€850 per dose, immunisation costs ranged from €2.4–€41.6 million (seasonal NmAb) and €4.9–€83.1 million (plus catch-up). Net investment increased sharply with dose cost, ranging from net savings to €31.8 million (seasonal NmAb) and up to €63.8 million (seasonal plus catch-up NmAb). RoI declined from 4.0 to 0.2 for both strategies as costs rose, reflecting diminishing returns. The seasonal NmAb strategy consistently showed higher RoI and lower net investment than the catch-up strategy across all cost levels.

Additional results of budget impact analysis by region are detailed in the KCE report, titled “Cost-effectiveness of new preventive options against RSV infections in Belgian infants [1].

*Table S. 16 – Treatment costs avoided, immunisation costs, return on investment and direct net benefits per year: MV at 40% coverage versus ‘no intervention’ from the HCP perspective (Mean [95%CrI])*

| | Treatment costs avoided (‘000 €) | | Immunisation costs (assumed all-inclusive*) (‘000 €) | | Net investment (‘000 €) | | Return on investment ratio (RoI) | |
|---|---|---|---|---|---|---|---|---|
| ***Cost per dose*** | MV | MV: Sep-Mar | MV | MV: Sep-Mar | MV | MV: Sep-Mar | MV | MV: Sep-Mar |
| ***Belgium (nr of doses)*** | | | (43,472) | (25,359) | | | | |
| *€50* | 5,102 [2,986 ; 7,271] | 4,189 [2,547 ; 5,683] | 2,174 | 1,268 | -2,928 [-5,097 ; -812] | -2,921 [-4,415 ; -1,280] | 2.35 | 3.30 |
| *€100* | | | 4,347 | 2,536 | -755 [-2,924 ; 1,361] | -1,653 [-3,147 ; -12] | 1.17 | 1.65 |
| *€150* | | | 6,521 | 3,804 | 1,419 [-750 ; 3,535] | -385 [-1,879 ; 1,256] | 0.78 | 1.10 |
| *€200* | | | 8,694 | 5,072 | 3,593 [1,423 ; 5,709] | 883 [-611 ; 2,524] | 0.59 | 0.83 |
| *€250* | | | 10,868 | 6,340 | 5,766 [3,597 ; 7,882] | 2,151 [657 ; 3,792] | 0.47 | 0.66 |

*HCP: health care payers’, MV: year-round single-dose maternal vaccine (MV) during pregnancy, MV: Sep-Mar: seasonal maternal vaccine from September to March, nr: number. * These vaccination costs should cover purchase, stockage, distribution and administration of the listed number of doses. Currently none of these cost items are fully known.*

*Table S. 17 – Treatment costs avoided, immunisation costs, return on investment and direct net benefits per year: NmAb at 90% coverage versus ‘no intervention’ from the HCP perspective (Mean [95%CrI])*

| | Treatment costs avoided (‘000 €) | | Immunisation costs (assumed all-inclusive*) (‘000 €) | | Net investment (‘000 €) | | Return on investment ratio (RoI) | |
|---|---|---|---|---|---|---|---|---|
| ***Cost per dose*** | NmAb: Oct-Mar | NmAb: Oct-Mar + catch-up | NmAb: Oct-Mar | NmAb: Oct-Mar + catch-up | NmAb: Oct-Mar | NmAb: Oct-Mar + catch-up | NmAb: Oct-Mar | NmAb: Oct-Mar + catch-up |
| ***Belgium (nr of doses)*** | | | (48,906) | (97,812) | | | | |
| *€50* | 9,772 [8,638 ; 10,689] | 19,351 [17,013 ; 21,315] | 2,445 | 4,891 | -7,326 [-8,244 ; -6,193] | -14,460 [-16,424 ; -12,122] | 4.00 | 3.96 |
| *€150* | | | 7,336 | 14,672 | -2,436 [-3,353 ; -1,302] | -4,679 [-6,643 ; -2,341] | 1.33 | 1.32 |
| *€250* | | | 12,226 | 24,453 | 2,455 [1,537 ; 3,588] | 5,102 [3,138 ; 7,440] | 0.80 | 0.79 |
| *€350* | | | 17,117 | 34,234 | 7,345 [6,428 ; 8,479] | 14,883 [12,919 ; 17,221] | 0.57 | 0.57 |
| *€450* | | | 22,008 | 44,015 | 12,236 [11,318 ; 13,369] | 24,664 [22,700 ; 27,002] | 0.44 | 0.44 |
| *€550* | | | 26,898 | 53,797 | 17,127 [16,209 ; 18,260] | 34,446 [32,482 ; 36,784] | 0.36 | 0.36 |
| *€650* | | | 31,789 | 63,578 | 22,017 [21,100 ; 23,150] | 44,227 [42,263 ; 46,565] | 0.31 | 0.30 |
| *€750* | | | 36,680 | 73,359 | 26,908 [25,990 ; 28,041] | 54,008 [52,044 ; 56,346] | 0.27 | 0.26 |
| *€850* | | | 41,570 | 83,140 | 31,798 [30,881 ; 32,932] | 63,789 [61,825 ; 66,127] | 0.24 | 0.23 |

*HCP: health care payers’, NmAb: Oct-Mar: seasonal NmAb strategy from October to March, NmAb: Oct-Mar + catch-up: seasonal NmAb combined with a catch-up strategy, nr: number. * These vaccination costs should cover purchase, stockage, distribution and administration of the listed number of doses. Currently none of these cost items are fully known.*
